# Circulating microRNAs are related to cognitive domains in the general population

**DOI:** 10.1101/2024.05.07.24306994

**Authors:** Konstantinos Melas, Valentina Talevi, Rika Etteldorf, Santiago Estrada, Dennis M. Krüger, Tonatiuh Pena, N. Ahmad Aziz, André Fischer, Monique M.B. Breteler

## Abstract

**INTRODUCTION:** Circulating microRNAs have been suggested as candidates for detecting and preventing subclinical cognitive dysfunction. However, replication of previous findings and identification of novel microRNAs associated with cognitive domains, including their relation to brain structure and the pathways they regulate, are still lacking.

**METHODS:** We examined circulating microRNAs and microRNA *co-expression* clusters in relation to cognitive domains, structural MRI measures, and target gene expression in 2869 participants of a population-based cohort.

**RESULTS:** Five previously identified and 10 novel microRNAs were associated with cognitive domains. Importantly, seven of these microRNAs were also associated with cortical thickness and two with hippocampal volume. Functional genomics analysis showed that the identified microRNAs regulated genes in pathways like neurogenesis, axon guidance, and synapse assembly.

**DISCUSSION:** We identified microRNAs associated with cognitive domains, brain regions, and neuronal processes affected by aging and neurodegeneration, making them promising candidate blood-based biomarkers or therapeutic targets of subclinical cognitive dysfunction.

## 1 BACKGROUND

Mitigating the impact of cognitive aging remains one of the biggest challenges of the 21^st^ century [1]. Early detection of subclinical cognitive dysfunction is necessary to enable interventions at the earliest stages of its pathophysiology [2]. Currently, tailored neuropsychological test batteries are the gold standard for detecting cognitive dysfunction. However, such batteries can be time-consuming, require experienced examiners, and are dependent on individual traits such as language and cultural background. Moreover, their sensitivity in the very early stages of cognitive dysfunction, and in highly educated persons, is limited [3]. The identification of blood-based molecular biomarkers of subclinical cognitive dysfunction could facilitate earlier and more reliable detection of individuals at risk of accelerated cognitive aging or neurodegeneration [4]. Additionally, such biomarkers might point to specific pathophysiologic processes involved in cognitive aging, and thus help the development of targeted interventions.

Various blood-based biomarkers have been investigated for the early detection of cognitive dysfunction, including neurodegenerative [4], lipidomic, inflammatory [5] and proteomic [6] markers. A highly promising line of research focuses on microRNAs [7,8]. These small, endogenous, non-coding RNAs bind to target genes, mainly through base pairing, leading to degradation of these genes or inhibition of their translation. A single microRNA can have several physiological effects through the regulation of multiple genes [9]. Additionally, microRNAs are highly expressed in the brain, where they regulate functions like synaptic plasticity and memory. Brain-enriched microRNAs can cross the blood-brain barrier and enter the peripheral circulation, where they remain stable for extended periods [8]. Thus, microRNAs have been investigated for the diagnosis and treatment of neurodegenerative diseases [10]. For example, circulating microRNA-based panels were shown to have high accuracy in diagnosing Alzheimer’s Disease (AD), and one such diagnostic panel has entered phase I of clinical development [11]. Conversely, microRNA-based therapeutics have been hindered by the incomplete characterization of the functions of tested microRNAs, which can lead to unforeseen side effects [11].

To assess whether circulating microRNAs can be employed for early detection or prevention of cognitive dysfunction, their association with cognition must be investigated in non-demented individuals, and their functions must be better understood. In the past, a few population-based studies identified microRNA signatures associated with general cognitive function and decline [12,13], verbal memory changes longitudinally [14], or multiple cognitive domains cross-sectionally [15]. A recent study identified a three-microRNA signature related to cognitive function and decline, by combining population-level and animal model data [16]. However, most of these studies had relatively small samples, potentially limiting their capability to detect microRNAs with weaker effect sizes. Specifically, the largest study included 1615 participants [12], while the rest had sample sizes ranging from 132 [16] to 830 [15] participants. Moreover, the only study that examined more than one cognitive domain evaluated only a small number (n=38) of microRNAs previously linked to AD [15]. Thus, there is a need for studies combining larger sample sizes, multiple cognitive domains, and the untargeted evaluation of many microRNA transcripts. In addition, it is worth noting that only five microRNAs (miR-4732-3p, miR-363-3p, miR-181a-5p, miR-146a-5p and let-7c-5p) overlapped among at least two of these studies. This reflects a greater problem in microRNA research. The large heterogeneity in microRNA sampling and quantification methods often leads to inconsistent results among studies, which highlights the need for replication [17]. Lastly, none of these studies examined the association of cognition-related microRNAs with brain structure or gene expression. This could provide information on key microRNA-regulated biological pathways, enabling their use as therapeutic targets against cognitive decline.

Here, we aimed to first identify blood-circulating microRNAs associated with the major domains of cognitive function, by replicating the results of previous studies and identifying novel associations in a population-based cohort consisting of 2869 individuals. Next, we determined which of the identified cognition-related microRNAs were associated with brain structure. Lastly, leveraging gene expression data through a functional genomics approach, we elucidated the molecular pathways through which the identified microRNAs could affect cognitive function and brain structure.

## 2 METHODS

### 2.1 Study Design

Our analysis was based on cross-sectional baseline data from the Rhineland Study, an ongoing prospective, population-based cohort study in Bonn, Germany. All residents of two geographically defined areas in Bonn, aged 30 years or older, were invited to participate in the study. The only exclusion criterion was an insufficient command of the German language to provide informed consent. One of the main goals of the Rhineland Study is to investigate biomarkers and determinants of aging and age-related diseases in the general population, following a deep-phenotyping approach. Each participant undergoes a comprehensive 7-hour examination protocol, which includes detailed questionnaires, brain imaging, neuropsychological examinations, and blood sample analyses.

### 2.2 Standard protocol approvals, registrations, and participant consent

Approval to undertake the study was obtained from the ethics committee of the University of Bonn, Medical Faculty. The study is carried out in accordance with the recommendations of the International Conference on Harmonization Good Clinical Practice standards. We obtained written informed consent from all participants in accordance with the Declaration of Helsinki.

### 2.3 Study population

Biomaterial was selected from the first 3000 participants of the Rhineland Study who provided blood samples for microRNA sequencing. MicroRNA expression data was missing due to technical issues (n=33) or exclusion during quality control (n=38). From the remaining 2929, we excluded 60 participants because of missing cognition data (due to: contraindications/exclusion criteria, n=7; participant refusal, n=3; technical issues, n=3; exclusion during quality control, n=9; a combination of the above, n=38), leaving 2869 participants with both microRNA expression and cognitive performance data (“cognition dataset”). Brain imaging data were available for a subset of 2045 participants of the cognition dataset (brain imaging data missing due to: contraindications/exclusion criteria, n=452; participant refusal, n=330; other/ unknown reasons, n=42). Gene expression and blood cell count data were available for a subset of 2138 participants of the cognition dataset (gene expression data missing due to: technical issues, n=475; exclusion during quality control, n=129; blood cell count data missing due to: technical issues, n=126; unknown reasons, n=1). The overlap of the subset with complete MRI data and the subset with complete gene expression and blood cell count data consisted of 1534 participants.

### 2.4 Blood sample acquisition and RNA isolation

Blood samples were collected between 7:00 to 9:45 in the morning from an antecubital or dorsal hand vein. On some occasions, blood sample collection was performed on a different visit than the cognitive evaluation. The time difference between the two visits was usually less than one month. Whole blood samples for microRNA and gene expression (messenger RNA) sequencing were collected in PAXgene Blood RNA tubes (PreAnalytix/Qiagen) and stored at -80° Celsius. Before sequencing, PAXgene Blood RNA Tubes were thawed and incubated at room temperature to increase RNA yield. Total RNA was isolated according to manufacturer’s instructions using the PAXgene Blood miRNA Kit and following the automated purification protocol (PreAnalytix/Qiagen).

### 2.5 MicroRNA and gene expression measurement

We performed the microRNA sequencing on the Illumina HiSeq 2000 over 44 batches, and the RNA sequencing on the NovaSeq6000 platform over 28 batches. Detailed information on our microRNA and gene sequencing and analysis pipeline is provided in **Supplementary Methods 1**. MicroRNAs and genes with overall mean expression levels greater than 15 reads and expressed in at least 5% of the participants were included for further analysis, resulting in a total of 415 microRNAs and 11,325 genes. Raw counts were normalized and transformed using the *varianceStabilizingTransformation* function from *DESeq2* (v1.30.1), which includes a log2 transformation [18]. Mean normalized counts of sequenced microRNAs ranged from 3.09 (for miR-4707-3p) to 21.34 (for miR-486-5p). As a last step, microRNA and gene expression data were adjusted for sequencing batch effects by creating a linear regression model, with microRNA normalized counts as the dependent variable and sequencing batch as the independent variable, and extracting the model residuals.

### 2.6 Assessment of cognitive function

The neuropsychological test battery of the Rhineland Study has been described in detail elsewhere [19]. Examined domains included working memory, episodic verbal memory, processing speed, executive function, and crystallized intelligence. Working memory was assessed with the orally performed Digit Span forward and backward task, and the touchpad-based Corsi block-tapping test, based on the Psychology Experiment Building Language battery [20]. Episodic verbal memory was evaluated with the Auditory Verbal Learning and Memory test with a list length of 15 words [21]. Assessment of processing speed was based on a numbers-only trail-making test (Trail-making test A), and prosaccade latency (time needed to initiate a saccade), derived from an eye movement test battery [22]. We examined executive function using the antisaccade error rate (percentage of trials in which the participant made a direction error) from the same battery, combined with a categorical word fluency task (animals), and number-and-letters trail-making test (Trail making test B). Lastly, we measured crystallized intelligence with the 37-item Mehrfachwahl-Wortschatz-Intelligenztest [23]. Cognitive domain performance scores were created by averaging the z-scores of the tests contributing to each domain. The creation of these z-scores was based on data from the first 5000 participants of the Rhineland Study. Additionally, we averaged the z-scores for working memory, episodic verbal memory, processing speed, and executive function to create a global cognition composite score. All examinations were administered in the German language, following a standardized procedure by certified study technicians.

### 2.7 Image acquisition and brain segmentation

Participants underwent a one-hour examination protocol in 3T MRI scanners (Siemens Prisma Magnetom, Erlangen, Germany) equipped with a 64-channel head-neck coil [24]. During the MRI section, a T1-weighted multi-echo magnetization prepared rapid gradient echo (MPRANGE) image was acquired [25]. T1-weighted images were processed with the standard Freesurfer (version 6.0) recon-all pipeline [26] to obtain the brain imaging phenotypes. For our analysis of MRI data, we evaluated regions relevant to cognitive aging and neurodegeneration, namely mean and regional cortical thickness, and hippocampal volume [27]. In addition, we evaluated total brain volume as a general indicator of brain health. Regional cortical thickness was evaluated for the 31 cortical Regions of Interest (ROIs) included in the Desikan–Killiany–Tourville (DKT) cortical atlas [28], and was separately assessed for the left and right hemispheres.

### 2.8 Demographic and biochemical variables

Participants’ age and sex were included as demographic variables. Information on educational level, native language, and history of physician-diagnosed neuropsychiatric or neurodegenerative disorders was based on self-reports. Educational level was determined using the International Standard Classification of Education 2011 and was coded as low (lower secondary education or below), middle (upper secondary education to undergraduate university level), and high (postgraduate university study). Native language was coded as German or other. Differential blood cell count measurements (erythrocytes, nucleated erythrocytes, leukocytes, and platelets) were performed at the Central Laboratory of the University Hospital in Bonn, using EDTA-whole blood samples on a hematological analyzer Sysmex XN9000.

### 2.9 Statistical analysis

Descriptive data were expressed as mean ± standard deviation (SD) and counts with proportions for numerical and categorical variables. To compare cognitive scores among the main analytical sample and the subsamples used for brain imaging and gene expression analyses, we used Type III Analysis of Covariance adjusting for age and sex.

To examine the association of microRNA expression with cognitive scores, we followed two complementary methods. First, based on the *guilt-by-association* framework [29], we assumed that microRNAs that follow similar expression patterns act jointly to form biologically functional units. We utilized the Weighted Gene Co-expression Network Analysis (WGCNA) method [30], to create groups of similarly expressed microRNAs (*co-expression* analysis). This allowed us to account for the correlated structure of microRNA expression data, increasing our statistical power for detecting trait-related microRNAs. Second, we performed a *per microRNA* analysis, meaning that we directly examined the association of individual microRNAs with cognitive scores. This was done to account for single microRNAs that might be strongly related to cognitive outcomes, but only weakly co-expressed with other microRNAs, in which case the grouping method would be less effective.

#### 2.9.1 Construction of weighted gene co-expression network

We clustered microRNAs in *co-expression* modules using the *WGCNA* R package (v.1.70-3) [30,31]. We chose a soft thresholding power of 7 based on scale-free topology, and we used the Hybrid Dynamic Tree Cut method to identify microRNA modules, with a minimum module size of 5. This resulted in 16 modules, which we refer to by randomly assigned colors (green, magenta, turquoise, black, pink, yellow, brown, red, blue, greenyellow, purple, cyan, salmon, lightcyan, midnightblue, tan). We then reduced the expression of all microRNAs in a module to a single value (module expression). Detailed information on the application of the WGCNA algorithm is provided in **Supplementary Methods 2.**

#### 2.9.2 Association of module and microRNA expression with cognition

For the *co-expression* and *per microRNA* analyses we examined the association of module and microRNA expression with cognitive scores using multiple linear regression. Firstly, we converted all numeric variables to z-scores, to permit the comparability of all linear regression coefficients. Then, for each module, each microRNA, and each cognitive score we created a separate model including module or microRNA expression as the independent variable and cognitive score as the dependent variable while adjusting for age and sex (Model 1: *cognitive score* ∼ *microRNA or module expression* + *age* + *sex).* Additionally, we created a second model that included educational level and native language as covariates. For crystallized intelligence, however, we did not adjust for native language as German as a native language is a prerequisite for undertaking the test (Model 2: *cognitive score* ∼ *microRNA or module expression* + *age* + *sex* + *educational level [+ native language]).* Lastly, we performed a sensitivity analysis by excluding participants who reported a physician-diagnosed neuropsychiatric or neurodegenerative disorder (i.e., dementia, Parkinson’s disease, multiple sclerosis, stroke, schizophrenia), and re-ran Model 1 in the remaining sample.

#### 2.9.2.1 Replication of microRNAs previously associated with cognition and AD or MCI

The first part of our *per microRNA* analysis consisted of a replication analysis for *a priori* selected microRNAs. This included 46 microRNAs associated with cognition in previous similar studies [12–16], and 10 microRNAs consistently found to be differentially expressed in the blood of AD and Mild Cognitive Impairment (MCI) patients in a recent meta-analysis [17]. An overview of these studies can be found in **Supplementary Table S1**. As all of these studies only examined measures of fluid intelligence, for this analysis we only included cognitive scores that reflect fluid intelligence (i.e., global cognition, working memory, episodic verbal memory, processing speed, and executive function). One study examined the association of microRNA expression with cognition using linear regression, as well as comparing expression between two cognitive performance categories [15]. For this study, we included for replication microRNAs from the linear regression analysis, which presented greater similarity with our own method. *P* values for the replication analysis were not corrected for multiple testing.

##### 2.9.2.2 Hypothesis-free analysis

Beyond the replication analysis, the association of microRNAs with cognition was evaluated without *a priori* hypotheses and thus we corrected for multiple testing using the Benjamini-Hochberg false discovery rate (fdr) method. Specifically, *P* values were corrected for the number of modules (n=16) for the *co-expression* analysis, and for the number of microRNAs (n=415) for the *per microRNA* analysis.

After identifying modules significantly associated with cognitive scores in the *co-expression analysis*, we also identified hub microRNAs in these modules. These were defined as microRNAs significantly associated with the same cognitive score as the module, and highly correlated with module expression (*high significance* and *high membership* microRNAs; **Supplementary Methods 2**). Here, *P* values were not corrected for multiple testing, as this was only done for the modules already significantly associated with cognitive scores. In all cases, the statistical significance level was set at 0.05 (fdr ≤ .05 or *P* value ≤ .05).

#### 2.9.3 Interaction and stratification based on age and sex

To examine whether age and sex modified the association of microRNAs with cognitive scores, we used additional linear models, including interaction terms between microRNA expression and age (Model 3: *cognitive score* ∼ *microRNA expression* + *age* + *microRNA×age* + *sex*), or between microRNA expression and sex (Model 4: *cognitive score* ∼ *microRNA expression* + *age* + *sex* + *microRNA×sex*). For microRNAs with a significant interaction term with either age or sex, we additionally performed a stratification analysis. This was done by splitting our main analytical sample based on median age in older (≥ 54 years) or younger (<54 years) participants, and in men or women. We then ran Model 1 again in these age and sex strata.

#### 2.9.4 Association of cognition-related microRNAs with brain MRI measures

MicroRNAs identified as significantly associated with cognition through the replication and hypothesis-free analyses were evaluated for their association with hippocampal volume, mean cortical thickness and total brain volume. MicroRNAs that were significantly associated with mean cortical thickness were further evaluated for their association with cortical ROI thickness. As for the association of microRNAs with cognitive scores, this was done by first converting numeric variables to z-scores and then employing multivariable linear regression. Brain *MRI* traits were included as the dependent variable and microRNA expression as the independent variable while adjusting for age and sex. When the dependent variable was hippocampal or total brain volume, we additionally adjusted for estimated Total Intracranial Volume (eTIV), to account for differences in head size (Model: brain *MRI measure* ∼ *microRNA or module expression* + *age* + *sex* [+ *eTIV*]). The results of this analysis were not corrected for multiple testing based on the *a priori* hypothesis that microRNAs associated with cognition would also be associated with brain structure.

### 2.10 MicroRNA expression in tissues

After identifying the microRNAs significantly associated with cognition, we assessed in which tissues they were most highly expressed. For this, we used miRNATissueAtlas2, an online resource that provides data on microRNA expression in 60 human tissues, acquired post-mortem from 6 donors [32]. Using this data, we converted microRNA expression across tissues to a z-score and we calculated the median of microRNA expression for each tissue across donors.

### 2.11 Functional genomics analysis

To determine genes that could be regulated by microRNA modules *in vivo*, we first obtained putative target genes for each microRNA using the *multimir* R package (v.1.12.0) [33]. Subsequently, we examined the association between microRNA and target gene expression in our partipants using linear regression, adjusting for age, sex, and blood cell counts (Model: *target gene expression* ∼ *microRNA expression* + *age* + *sex* + *erythrocyte count* + *nucleated erythrocyte count* + *leukocyte count* + *platelet count*). We kept for further analysis genes whose expression was negatively associated with the expression of the targeting microRNA. These genes were then used for a pathway enrichment analysis, pooling target genes of microRNAs belonging to the same WGCNA module, and using the Gene Ontology: Biological Processes database and the “*clusterProfiler*” (v. 3.18.1) R Bioconductor package [34]. Semantically similar terms were collapsed *post hoc* using the *rrvgo* (v. 1.2.0) R Bioconductor package [35], setting a small similarity threshold of 0.6. For detailed information on the identification of target genes and functional enrichment, refer to **Supplementary Methods 3**. We additionally examined the enrichment of target genes among genes identified in a published Genome-Wide Association Study (GWAS) of general cognitive function. This study identified 709 cognition-related genes, and gene expression data was available for 385 of those genes in our data [36]. The enrichment analysis was done by examining whether the target genes of each microRNA were overrepresented among the genes identified by the GWAS using a hypergeometric test (“*hypeR*” R Bioconductor package, v.1.10.0). This analysis was performed for individual microRNAs and not per module, to identify the most relevant single microRNAs. The statistical significance level fir enrichment analyses was set at .05 (fdr ≤ .05 or *P* value ≤ .05).

All analyses were performed using the R programming environment (v. 4.1.0).

### 2.12 Data availability

The data from the Rhineland Study are not publicly available due to data protection regulations. Access to data can be provided to scientists in accordance with the Rhineland Study’s Data Use and Access Policy. Requests for additional information or to access to the Rhineland Study’s datasets can be send to.

## 3 RESULTS

### 3.1 Participant characteristics

In our main analytical sample (cognition dataset, n=2869 participants), the mean age was 55.01 years, (SD=14.13 years, range=30-95 years) and 56% of participants were women. The subset of participants with complete MRI data was slightly younger and had slightly higher cognitive domain scores (**Table 1**). However, when adjusted for age and sex, the cognitive domain scores did not significantly differ between the cognition dataset and its subsets (all *P* values > .1).

**Table 1:** Study participant characteristics.

| Variable | Participants with complete cognitive examination and microRNA expression data, N = 2869 <sup>*</sup> | Participants with complete cognitive examination, microRNA expression, and brain MRI data, N = 2045 <sup>†, ‡</sup> | Participants with complete cognitive examination, microRNA expression, and gene expression data, N = 2138 <sup>‡</sup> |
| --- | --- | --- | --- |
| Age (years), mean (SD) | 55.01(14.13) | 54.31(13.90) | 54.99 (14.18) |
| Sex, n (%) |  |  |  |
| Female | 1595 (56%) | 1149 (56%) | 1173 (55%) |
| Male | 1274 (44%) | 896 (44%) | 965 (45%) |
| Global cognition (z-score), mean (SD) <sup>¶</sup> | 0.01 (0.64) | 0.05 (0.62) | 0.01 (0.63) |
| Executive function (z-score), mean (SD) <sup>¶</sup> | -0.03 (0.75) | 0.02 (0.73) | -0.02 (0.75) |
| Working memory (z-score), mean (SD) <sup>¶</sup> | -0.01 (0.72) | 0.03 (0.71) | 0.00 (0.71) |
| Ep. verbal memory (z-score), mean (SD) <sup>¶</sup> | 0.06 (0.96) | 0.11 (0.93) | 0.04 (0.96) |
| Processing speed (z-score), mean (SD) <sup>¶</sup> | 0.02 (0.83) | 0.06 (0.82) | 0.03 (0.83) |
| Crystallized Intelligence (z-score), mean (SD) <sup>¶</sup> | 0.02 (0.96) | 0.06 (0.93) | 0.02 (0.96) |
| First language, n (%) |  |  |  |
| German | 2682 (94%) | 1914 (94%) | 2006 (94%) |
| Other | 182 (6.4%) | 130 (6.4%) | 129 (6.0%) |
| Educational level, n (%) |  |  |  |
| Low | 55 (1.9%) | 34 (1.7%) | 34 (1.6%) |
| Middle | 1248 (44%) | 844 (42%) | 928 (44%) |
| High | 1543 (54%) | 1152 (57%) | 1155 (55%) |
| Left hippocampal volume (mm<sup>3</sup>), mean (SD) | - | 3.89×10<sup>3</sup> (0.46×10<sup>3</sup>) | 3.88×10<sup>3</sup> (0.46×10<sup>3</sup>) |
| Right hippocampal volume (mm<sup>3</sup>), mean (SD) | - | 4.05 ×10<sup>3</sup> (0.49×10<sup>3</sup>) | 4.04 ×10<sup>3</sup> (0.49×10<sup>3</sup>) |
| Total brain volume (mm<sup>3</sup>), mean (SD) | - | 1116.40×10<sup>3</sup> (120.73×10<sup>3</sup>) | 1115.78×10<sup>3</sup> (120.02×10<sup>3</sup>) |
| eTIV(mm<sup>3</sup>), mean (SD) | - | 1.56×10<sup>6</sup> (0.16×10<sup>6</sup>) | 1.56×10<sup>6</sup> (0.16×10<sup>6</sup>) |
| Mean cortical thickness (mm), mean (SD) | - | 2.45 (0.09) | 2.45 (0.09) |

\* Participants with data additionally missing for crystallized intelligence: n=190
† Values have not been adjusted for head size. Participants with data additionally missing for individual imaging markers: cortical thickness: n=17; total brain volume: n=18; hippocampal volume: n=4
‡ Subset of participants with complete cognitive examination and microRNA expression data
¶ Cognitive examination z-scores were generated based on summary statistics from the first 5000 participants of the Rhineland Study.

### 3.2 Association of microRNAs and microRNA co-expression modules with cognition

#### 3.2.1 Replication of microRNAs previously associated with cognition, AD, or MCI

Among the 46 microRNAs associated with cognition in previous population-based studies, miR-340-5p, miR-125b-5p, mir-4732-3p and miR-92a-3p were associated with at least one cognitive score in the same direction in our *per microRNA* analysis. Of note, miR-4732-3p had been found in two of the previous studies [12,13] and, among these 46 microRNAs, it had the strongest association with cognition in our data. Conversely, seven microRNAs were significantly associated with cognitive scores in the opposite direction in our data, compared to previous studies. Six of these microRNAs, miR-30e-3p, miR-342-5p, miR-574-5p, miR-320c, miR-215-5p, miR-320b, miR-122-5p, came from the same study by Comfort et al [13], which had included older (median age: 71 years) and only male participants. The seventh microRNA, miR-122-5p, had been identified in a study by Yaqub et al [12], which also included older participants (median age: 70.3 years) compared to our study (**Figures 1A & 1B**). To examine whether these inconsistent findings were due to the age or sex differences in the examined populations, we included the six microRNAs from Comfort et al in a sex-stratified analysis, and all seven microRNAs in an age-stratified analysis (shown in Section

**Figure 1:**
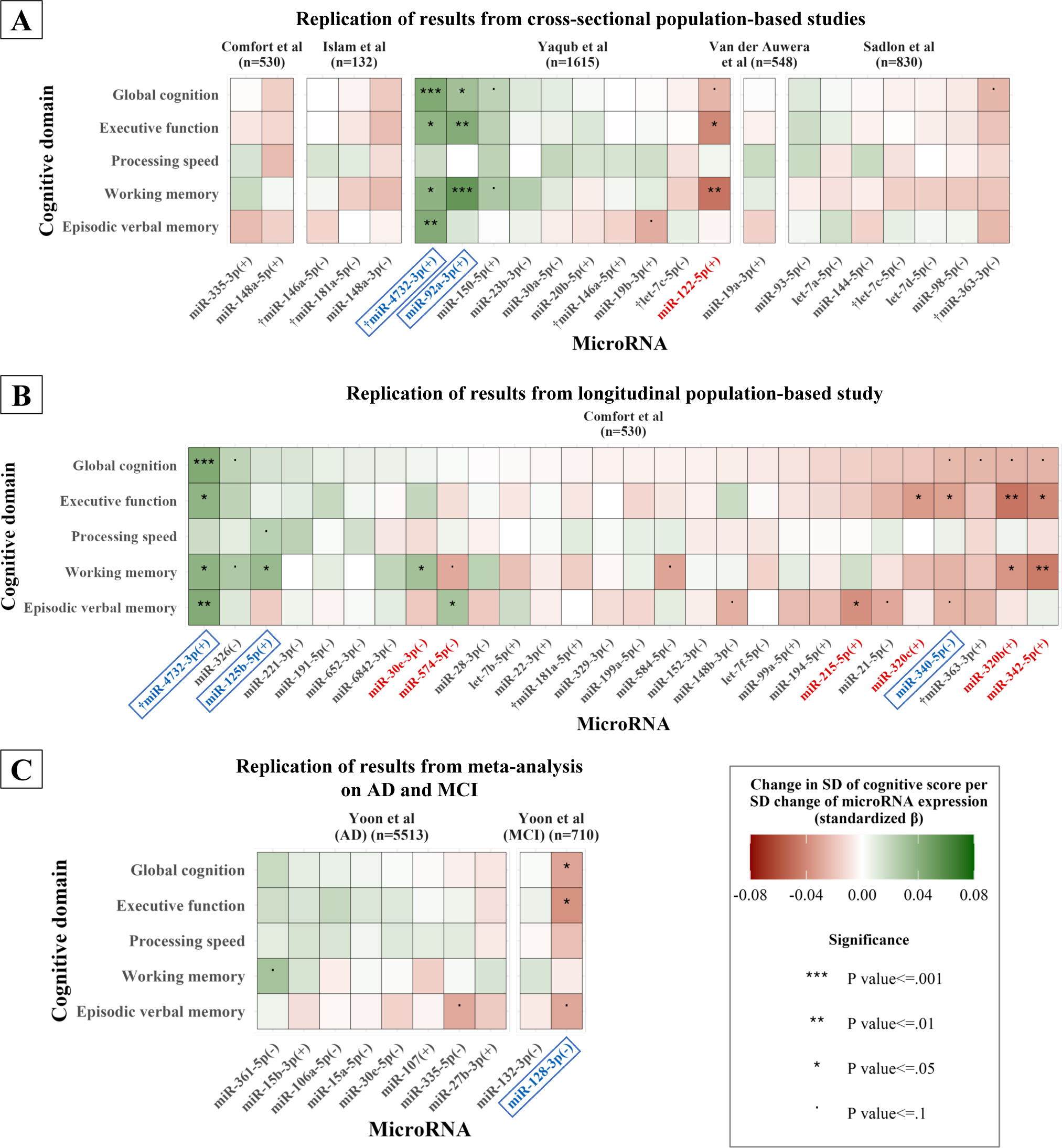
Replication of previous findings on microRNAs associated with cognition, AD or MCI. Replication of microRNAs that were identified as associated with cognition in previous cross-sectional population-based studies [12–16] **(A)**, one previous longitudinal population-based study **(B)** [13], and a previous meta-analysis of microRNAs dysregulated in AD and MCI **(C)** [17]. Details on the studies included can be found in **Supplementary Table S1.** The sample sizes of included studies are in parentheses next to study names. Statistical significance is indicated by the asterisks. MicroRNAs that appeared in multiple included studies are marked with “**†**”. MicroRNAs are marked with “+” in parentheses when their higher expression was associated with better cognitive function in included studies, and with “-” in the opposite case. For the AD and MCI meta-analysis, “+” indicates higher expression of the microRNA in controls compared to patients. MicroRNAs that were significantly associated with cognition with a similar direction in the included studies and in our data have been annotated with blue color and are outlined with a blue box. Annotated with red color are microRNAs that were significantly associated with cognition in our study, but in an opposite direction compared to included studies. Abbreviations: AD, Alzheimer’s Disease; MCI, Mild Cognitive Impairment; SD, Standard Deviation

*3.2.5*).

Among the 10 microRNAs differentially expressed in AD and MCI [17], higher miR-128-3p expression had been previously found higher in MCI patients and was associated with worse global cognition, executive function and episodic verbal memory performance in our data (**Figure 1C**).

#### 3.2.2 Hypothesis-free co-expression analysis

Beyond the replication of previous studies, we performed a hypothesis-free analysis to identify novel microRNA co-expression modules and hub microRNAs associated with cognition. Using WGCNA, we identified 16 microRNA co-expression modules with a size ranging from 5 to 129 microRNAs (**Suppleementary Fig. S1**). Modules were named after randomly assigned colors Adjusting for age and sex, and after correction for multiple testing, higher expression of only one of these modules (lightcyan) was significantly associated with worse episodic verbal memory (standardized ß: -0.048, 95% Confidence Interval [CI]: -0.078 to -0.019, fdr-corrected *P* value: .021, adjusted R^2^: 0.35). Additionally, we observed trends towards association of lower expression of another module (greenyellow) and higher expression of three modules (salmon, turquoise and pink) with better performance across most scores. However, none of these associations were significant after multiple testing correction **(Figure 2A)**. When additionally adjusting for educational level and first language, increased expression of the greenyellow module became significantly associated with worse executive function (standardized ß: -0.045, 95% CI: -0.075 to -0.016, fdr-corrected *P* value: .043, adjusted R^2^: 0.36). With the same adjustment, higher expression of the lightcyan module remained significantly associated with worse episodic verbal memory, yet the association slightly weakened (standardized ß: -0.041, 95% CI: -0.070 to -0.012, fdr-corrected *P* value: .097, adjusted R^2^: 0.39; **Figure 2B**). Based on these findings, we decided to retain the greenyellow and lightcyan modules for further analyses, as they demonstrated the strongest and most consistent associations with cognitive function.

**Figure 2:**
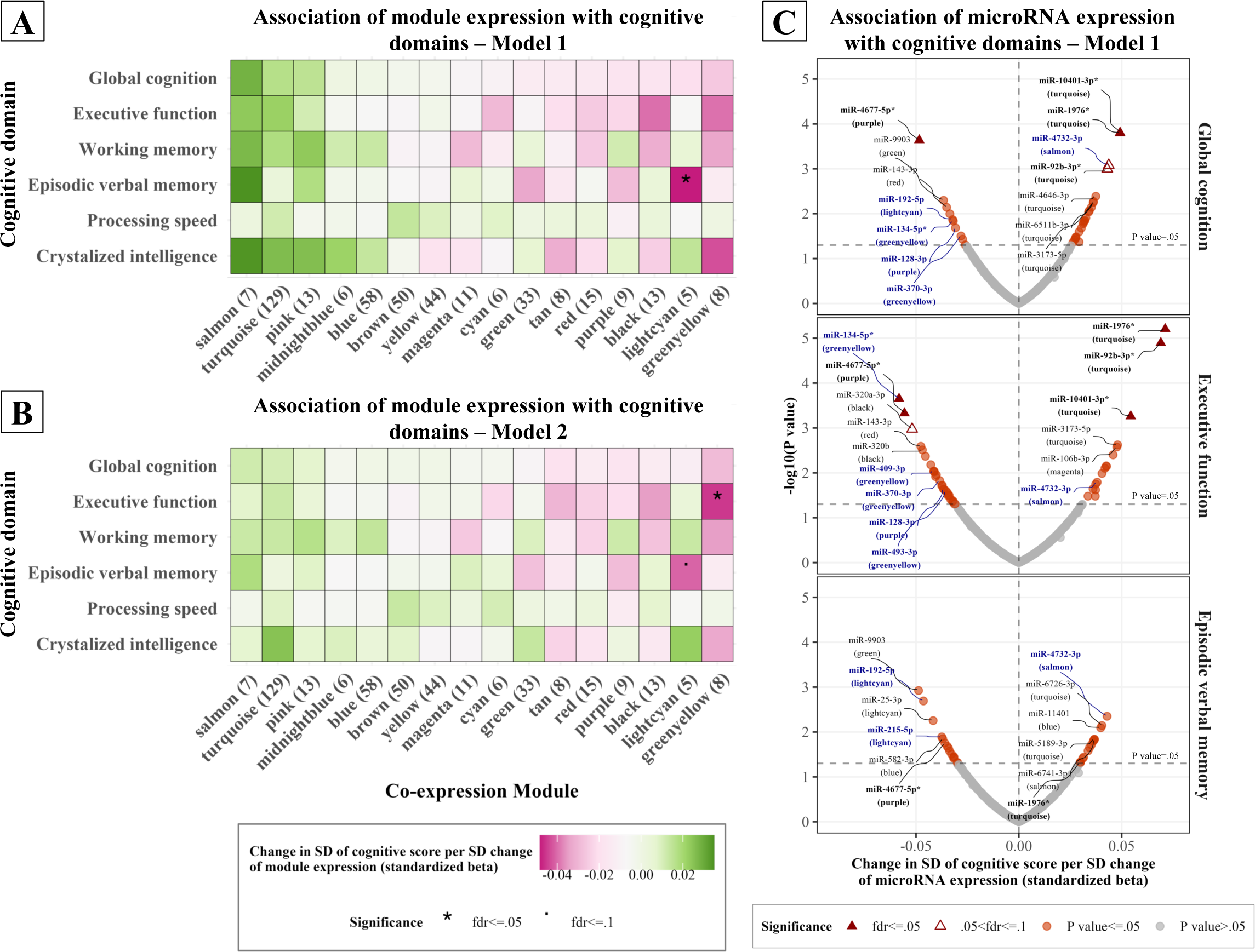
Hypothesis-free association of microRNA and module expression with cognition. **(A, B)** Linear regression coefficients and significance levels for the association between co-expression modules and cognitive scores. Shown are Model 1 (**A**) which was adjusted for age and sex, and Model 2 (**B**) which was additionally adjusted for first (native) language and educational level. Modules are named after colors, and numbers in parentheses indicate the number of microRNAs within each module. Significance was determined after correcting for multiple comparisons with the fdr method. **(C)** Volcano plots of the *per microRNA* association of all microRNAs measured in our study with global cognition, executive function and episodic verbal memory scores. The *P* value shown in the y-axis has not been corrected for multiple testing. Top microRNAs for each score are annotated and co-expression modules are shown in parentheses. MicroRNAs that reached statistical significance for at least one phenotype, after multiple testing correction (fdr ≤ 0.05), are shown with triangles and have been marked with an asterisk (*). Hub microRNAs are annotated with blue color. Abbreviations: fdr, false discovery rate; SD, Standard Deviation

We then identified hub microRNAs in these two modules, defined as microRNAs strongly correlated with module expression, while being significantly associated with the same cognitive domains as the modules (i.e., executive function for the greenyellow module and episodic verbal memory for the lightcyan module). In the model adjusted for age and sex, these conditions were fulfilled for four microRNAs from the greenyellow module, miR-134-5p, miR-409-3p, miR-370-3p and miR-493-3p, and for two microRNAs from the lightcyan module, miR-215-5p and miR-192-5p **(Figure 2C)**.

#### 3.2.3 Hypothesis-free per microRNA analysis

For the next step of the hypothesis-free analysis, we individually examined all microRNAs measured in our study (n=415) in relation to cognitive function, regardless of co-expression module. Adjusting for age and sex, we identified four microRNAs, miR-92b-3p, miR-1976, miR-10401-3p and miR-4677-5p, which were significantly associated with executive function after multiple testing correction. The same microRNAs were significantly associated with global cognition, with the exception of miR-92b-3p, which was associated with global cognition at borderline significance (**Figure 2C)**. Results changed only slightly when additionally adjusting for education and native language (**Supplementary Fig. S2**). Of note, miR-92b-3p is closely related to miR-92a-3p, which was identified in our replication analysis (Section *3.2.1*). These two microRNAs belong to the same family (mir-25 family) and share highly similar mature sequences [37]. Despite that, their expression levels were only moderately correlated in our data (Pearson’s correlation coefficient: 0.37, 95% CI: 0.34 to 0.40) Thus, combining the results from our literature replication, as well as our *co-expression* and *per microRNA* hypothesis-free analyses, we detected a signature of five previously identified (miR-340-5p, miR-125b-5p, mir-4732-3p, miR-92a-3p, miR-128-3p) and 10 novel microRNAs (miR-192-5p, miR-134-5p, miR-370-3p, miR-409-3p, miR-493-3p, miR-215-5p, miR-4677-5p, miR-10401-3p, miR-1976, miR-92b-3p) that were strongly related to cognition. Notably, while significant associations were primarily found with executive function or episodic verbal memory scores, these 15 microRNAs were nominally associated with most other cognitive scores as well **(Figure 3A**). Median values and distribution of the expression (normalized counts) of the 15 microRNAs can be found in **Supplementary Fig. S3.**

**Figure 3:**
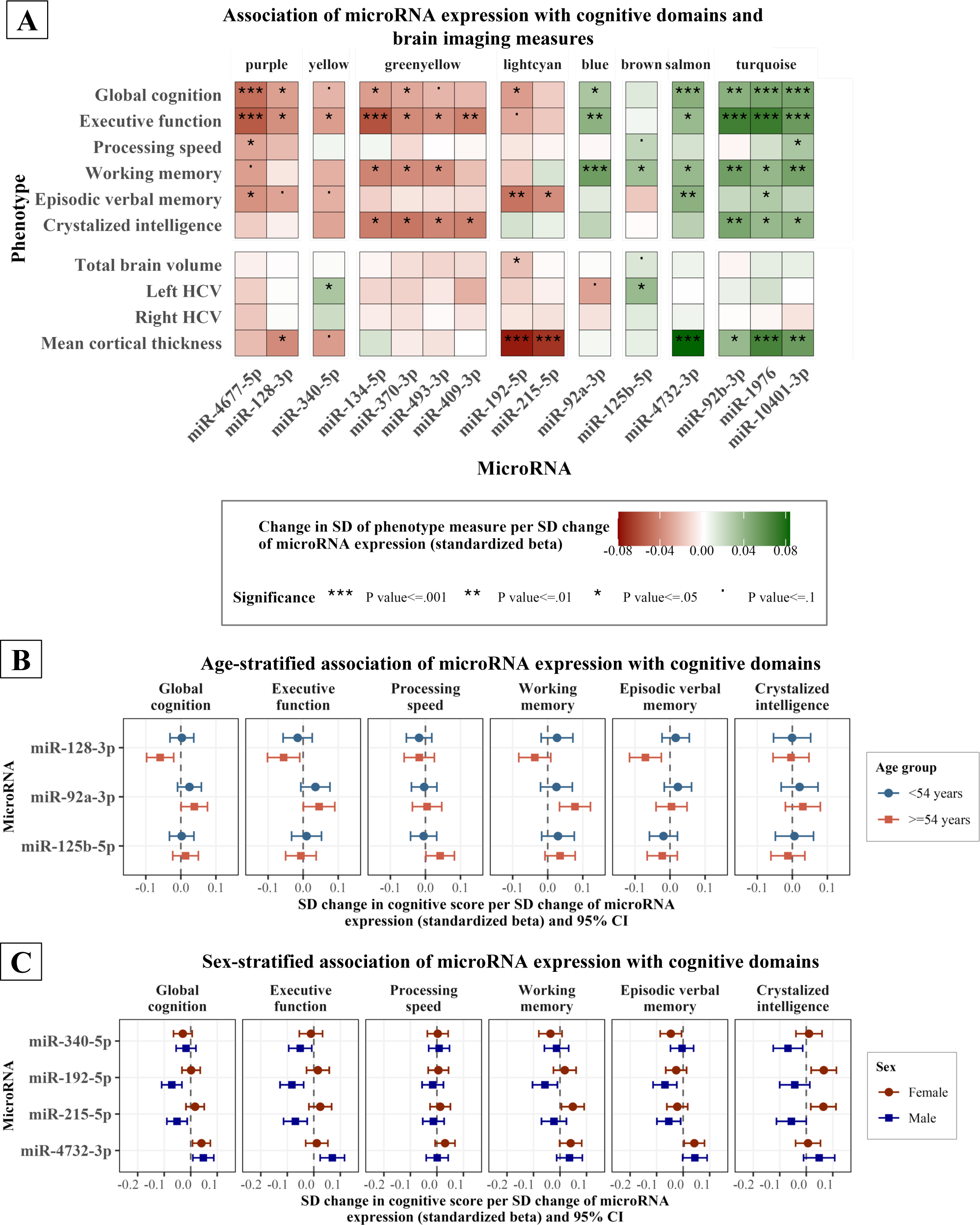
Association of microRNA expression with cognitive scores and brain imaging measures. The heatmap **(A)** shows the association of microRNA expression with cognitive scores and brain imaging measures. Included were microRNAs identified as associated with cognition in our literature replication, *co-expression* and *per microRNA* analyses. The association of some microRNAs with cognitive domain scores was modified by sex and age. This is illustrated in (**B**) for sex and **(C)** for age, which show the standardized regression betas and 95% confidence intervals for the sex- and age-stratified association of these microRNAs with each cognitive score. Linear regression analyses were based on Model 1, which included as covariates age and sex, except for the sex-stratified analysis which was controlled only for age. Abbreviations: SD, Standard Deviation; fdr, false discovery rate; HCV, Hippocampal Volume; CI, Confidence Interval indicates the standardized β for the regions in which microRNA expression was significantly associated with cortical thickness. The analysis was performed only for microRNAs that were significantly associated with cognition and mean cortical thickness across both hemispheres. The visualization was created with the *ggseg* (v. 1.6.5) and *ggsegDKT* (v. 1.0.1.1) R packages [75]. Abbreviations: SD, Standard Deviation

#### 3.2.4 Neuropsychiatric and neurodegenerative disorder sensitivity analysis

To ensure that our results were not confounded by dysregulation of microRNA expression due to neuropsychiatric or neurodegenerative disorders, we performed the same analysis after excluding participants with a self-reported physician diagnosis of dementia (n=3), Parkinson’s disease (n=8), multiple sclerosis (n=13), stroke (n=48), and schizophrenia (n=2). This sensitivity analysis resulted in only slight differences in linear regression coefficients for the 16 co-expression modules and the 15 cognition-related microRNAs, without affecting statistical significance (**Supplementary Fig. S4**).

#### 3.2.5 Age and sex as effect modifiers

Next, we examined whether the association of the microRNAs with cognition was modified by age or sex. We detected significant positive interaction terms with age for the associations of miR-128-5p with episodic verbal memory and global cognition, miR-125b-5p with executive function and crystallized intelligence, and miR-92a-3p with working memory. Age-stratified analysis confirmed that the association of these microRNAs with cognitive scores was stronger in the older age group (≥54 years; **Figure 3B**). Additionally, a significant sex interaction was observed for the association of miR-192-5p and miR-215-5p (lightcyan module) with multiple cognitive domains, miR-4732-3p with executive function and miR-340-5p with crystallized intelligence. Sex-stratified analysis showed that these microRNAs were more strongly associated with most cognitive domains in men. The effect modification was especially pronounced for miR-215-5p. Higher expression of this microRNA was associated with worse working memory and crystallized intelligence performance in the total sample and among men, but with better performance in these domains among women **(Figure 3C**).

Seven microRNAs, including the abovementioned miR-215-5p, were inconsistently associated with cognition in our study and in previous studies by Comfort et al. and Yaqub et al. [12,13] (Section *3.2.1*). As these previous studies included older participants compared to ours, the inconsistent associations of microRNAs with cognition might have been driven by age-dependent effects. Age-stratified analysis for these microRNAs partially confirmed this assumption. Specifically, for four microRNAs (miR-342-5p, miR-320b, miR-320c, miR-574-5p), the inconsistent association with cognition was stronger in younger participants and tended towards the null in older participants (**Supplementary Fig. S5A**). Similarly, the study by Comfort et al. included only male participants. For four of the microRNAs identified in this study (miR-342-5p, miR-320b, miR-320c, miR-30e-3p) the inconsistent association with cognition was stronger in women and tended towards the null in men (**Supplementary Fig. S5B**). However, the assumption of sex driving the inconsistent association with cognition was not confirmed for miR-215-5p. As mentioned above, higher miR-215-5p expression was associated with much worse cognitive function among men in our study, but with better cognitive function among the male population of Comfort et al.

### 3.3 Association of cognition-related microRNAs with brain MRI measures

Next, we assessed whether the expression of the 15 microRNAs identified as associated with cognition was also associated with mean cortical thickness, hippocampal volume, and total brain volume. Higher expression of miR-128-3p, miR-215-5p and miR-192-5p was significantly associated with a thinner cortex, and higher miR-192-5p expression was additionally associated with smaller total brain volume. On the contrary, higher expression of miR-4732-3p, miR-92b-3p, miR-1976 and miR-10401-3p was associated with a thicker cortex. Moreover, higher miR-125b-5p expression was associated with larger left hippocampal volume, which was consistent with its association with better cognitive function. Surprisingly, although miR-340-5p expression was associated with worse cognitive function, it was also associated with larger left hippocampal volume and, at borderline significance, a thinner cortex **(Figure 3A**).

ROI-specific analysis showed that the association of microRNAs with cortical thickness was overall strongest in the left hemisphere, and followed similar patterns for microRNAs in the same module. Specifically, the associations were strongest in the left middle temporal and postcentral gyri for lightcyan module microRNAs (miR-215-5p, miR-192-5p), and in the left precentral and superior temporal gyri for turquoise model microRNAs (miR-10401-3p, miR-1976, miR-92b-3p). For miR-4732-3p (salmon module), the associations were strongest bilaterally in the precentral and superior temporal regions, and in the right paracentral region. The association of miR-128-3p (purple module) with cortical thickness was overall weaker, and localized in the right middle and inferior temporal regions (**Figure 4**).

**Figure 4:**
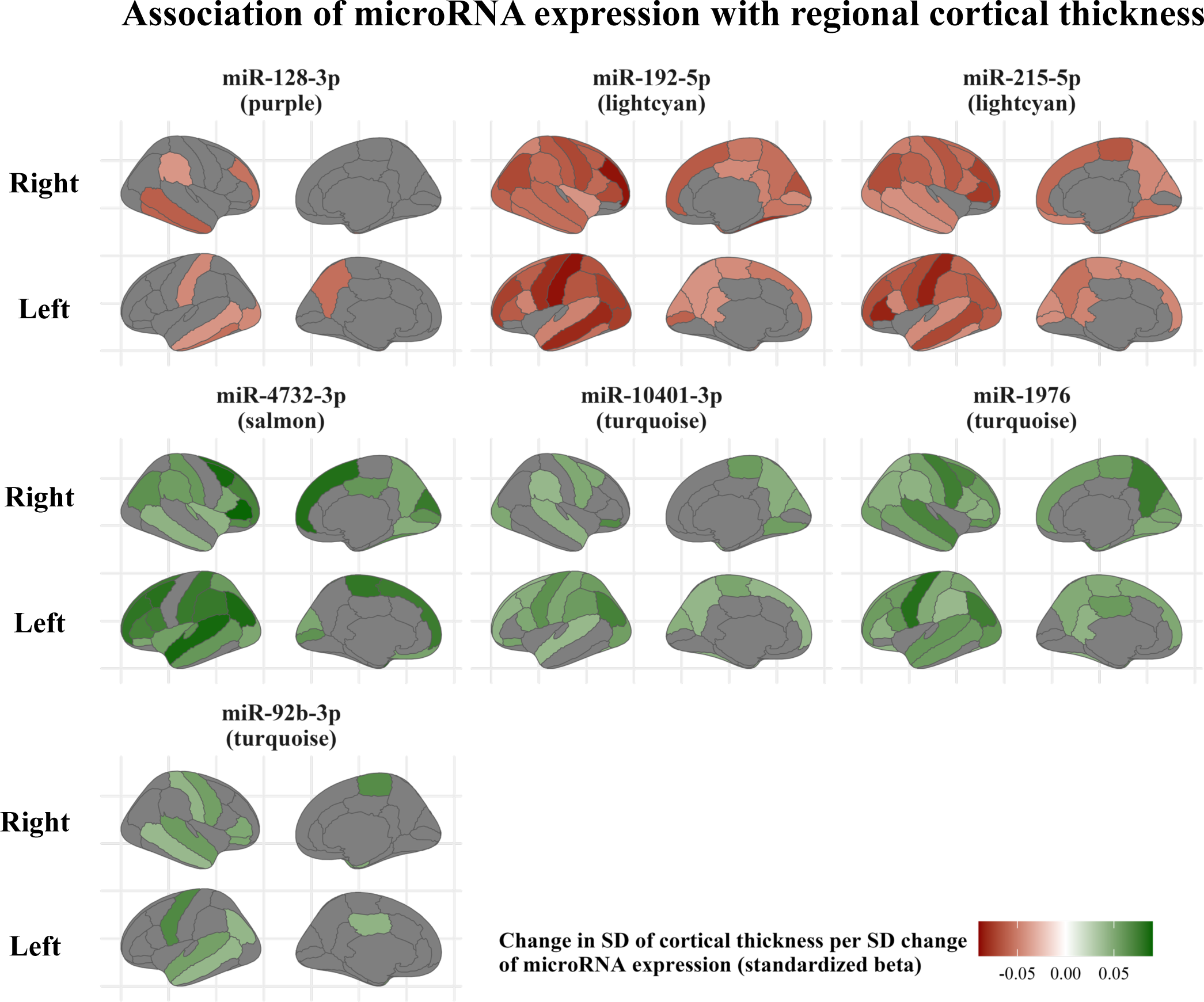
Region-specific association between microRNA expression and brain cortical thickness. Regions of interest have been mapped based on the Desikan-Killiany cortical atlas. The color indicates the standardized β for the regions in which microRNA expression was significantly associated with cortical thickness. The analysis was performed only for microRNAs that were significantly associated with cognition and mean cortical thickness across both hemispheres. The visualization was created with the *ggseg* (v. 1.6.5) and *ggsegDKT* (v. 1.0.1.1) R packages [75]. Abbreviations: SD, Standard Deviation

Lastly, it is worth mentioning that expression of miR-1976, miR-92b-3p, miR-128-3p and miR-192-5p was significantly or borderline significantly associated with the thickness of the entorhinal cortex.

### 3.4 MicroRNA expression in tissues

Using microRNATissueAtlas data, we determined that the majority of the 15 cognition-related microRNAs were expressed most abundantly across brain areas, such as the hippocampus, medulla oblongata, cerebellum, and cortex. MicroRNAs from the lightcyan module were the only exception to this, as they were expressed most abundantly in gastrointestinal tract tissues. The localization of miR-92b-3p and miR-128-3p in the cortex, hinted by their association with cortical thickness, was further reinforced by the high expression of these microRNA in the frontal lobe. Similarly, miR-125b-5p, which was associated with hippocampal volume, was also highly expressed in the hippocampus (**Figure 5**).

**Figure 5:**
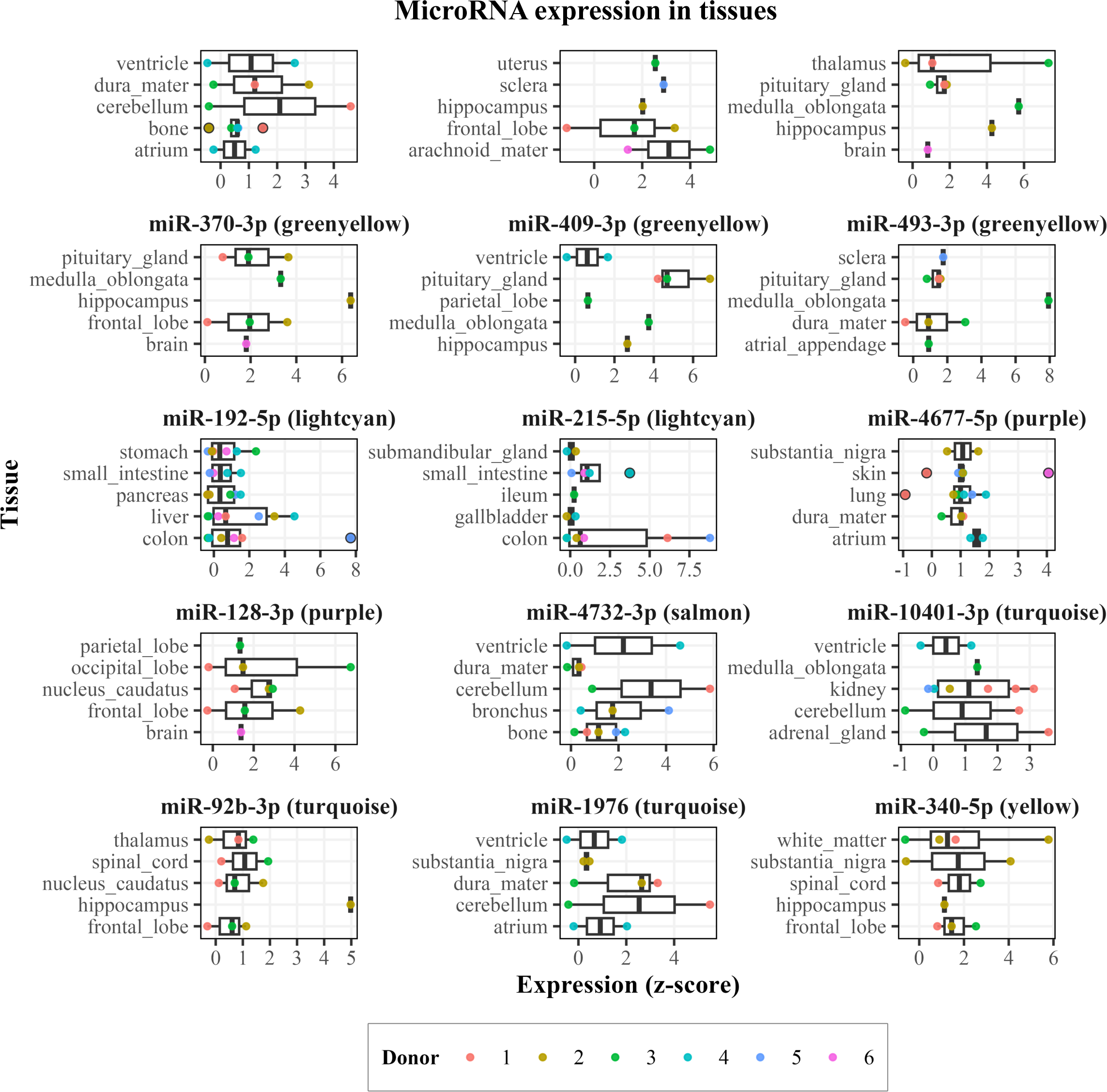
Tissue expression analysis of cognition-related microRNAs. Boxplots of tissue expression of microRNAs associated with cognition. Data was obtained from the microRNA Tissue Atlas [32], based on post-mortem samples taken from 6 human donors (here indicated by the colored dots). The module each microRNA belongs to is shown in parentheses. The top 5 tissues for each microRNA, based on median microRNA expression, are shown. Note that, to allow for better comparison of relative expression in tissues for each microRNA, the scale of the x-axis (expression z-score) varies.

### 3.5 Functional genomics analysis

#### 3.5.1 Identification of microRNA target genes

To better understand the function of the cognition-related microRNAs, we leveraged gene expression data, available in a subset of our study participants. In total, the 15 cognition-related microRNAs had 9072 putative target genes, and for 2560 of them expression of the targeting microRNA was negatively associated with gene expression. The close biological relation between miR-92a-3p and miR-92b-3p was corroborated by their relatively high number of shared negatively associated target genes (n=382 genes). Notably, these two microRNAs also shared a high number of negatively associated target genes with miR-128-3p (n=97). Beyond that, the 15 microRNAs had relatively few shared target genes (**Supplementary Fig. S6**).

#### 3.5.2 Pathway enrichment analysis of microRNA target genes

We then performed a Gene Ontology functional enrichment analysis of the negatively associated target genes per WGCNA module. This highlighted the distinct biological functions performed by each module. For example, the greenyellow module was highly enriched for “mitochondrial membrane permeability” and “mitochondrial transport”, the lightcyan module was highly enriched for “Wnt signaling”, “cell growth” and “wound healing”, while the turquoise module was highly enriched for “brain development”. Notably, pathways highly relevant to cognition were jointly enriched for several modules. This included “axon guidance”, “dendrite development”, “neurogenesis” and “synapse assembly” for the turquoise, purple, lightcyan and blue modules. Similarly, “transforming growth factor beta (TGF-β) signaling” was among the top results for the turquoise, brown, purple, lightcyan and blue modules, while “memory” was enriched for the turquoise, purple, greenyellow and blue modules. Moreover, “response to estradiol” was enriched for the lightcyan module **(Figure 6A).**

**Figure 6:**
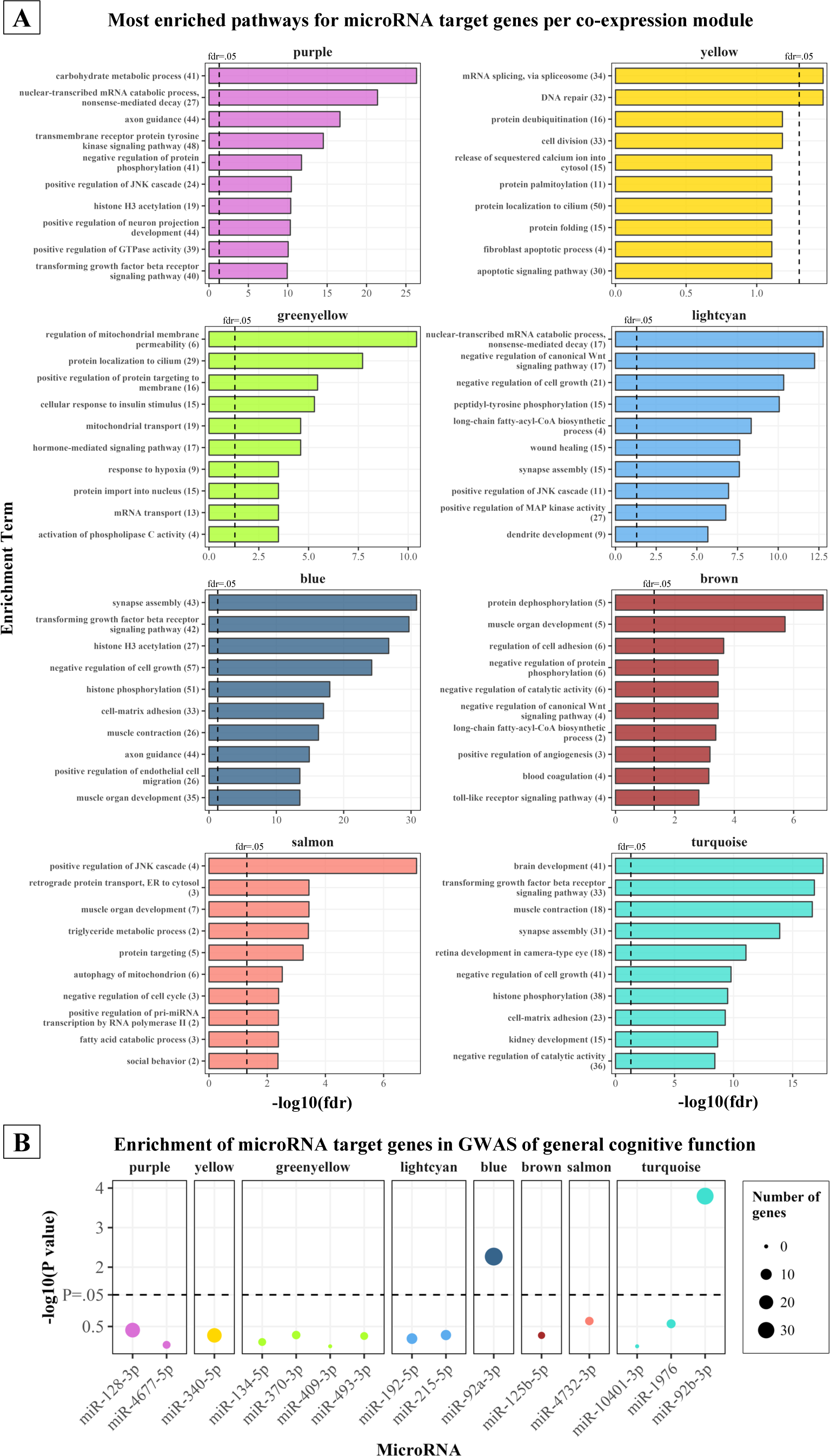
Function genomics analysis of cognition-related microRNAs (A) Results of *Gene Ontology: Biological Processes* enrichment analysis of negatively associated target genes of the microRNAs in each co-expression module. The top 10 most enriched terms (based on *P* values) are shown. The numbers in parentheses indicate the number of target genes in each enriched term. *P* values have been corrected for multiple testing using the fdr method. Note that, to allow for better comparison of relative enrichment in each module, the scale of the x-axis (negative logarithm of fdr) varies. (**B**) Overlap and overrepresentation (determined with a hypergeometric test) of negatively associated target genes of microRNA in a GWAS of global cognitive

#### 3.5.3 MicroRNA target genes in relation to general cognitive function

To further validate the involvement of the 15 identified microRNAs in cognition, we tested the overlap between microRNA target genes and genes identified by a recent GWAS of general cognitive function. Notably, only targets of miR-92b-3p and miR-92a-3p were significantly overrepresented among the genes identified by the GWAS **(Figure 6B**).

## 4 DISCUSSION

In the present study, we identified 15 microRNAs associated with cognitive domains in a large, general population-based cohort with a wide age range. Importantly, five of these microRNAs were identified by reproducing the results of previous studies. Our subsequent analyses using brain imaging and gene expression data enabled a detailed characterization of identified microRNAs, providing information on the mechanisms underlying their relation to cognition. Some microRNAs were associated with cortical thickness and hippocampal volume, while most were highly expressed in brain tissues and regulated key neuronal processes. Taken together, our large sample size, inclusion of previous studies, and analysis of multimodal data increase the robustness of our findings. Additionally, our results demonstrate the potential of identified microRNAs as biomarkers or therapeutic targets against cognitive dysfunction.

MicroRNAs associated with different cognitive domains or cortical regions could be investigated as blood-based biomarkers for disorders that preferentially affect these domains or regions, such as AD for memory and the temporal lobe, or Frontotemporal Dementia for executive function and the frontal lobe [38]. For example, miR-128-3p, which was upregulated in MCI patients in a past meta-analysis, was also associated with worse cognition in our study. In addition to MCI, miR-128-3p has also been dysregulated in AD [17], while suppressing its expression has been found to protect against AD pathology and memory loss in mice [39]. Moreover, we showed that it was associated with thickness in the temporal lobe and entorhinal cortex, which are affected very early in the pathophysiology of AD [40]. Early upregulation of this microRNA could indicate accelerated neurodegeneration, leading to brain atrophy and cognitive dysfunction. This would make it an ideal target for detecting and preventing AD-related cognitive dysfunction. However, contrary to the findings of our and other studies, a previous population-based study found that miR-128-3p was downregulated in participants with worse cognition [15]. Thus, further investigations are needed to determine its exact association with cognition.

Several of the identified microRNAs were associated with cortical thickness in regions involved in various cognitive processes. This corroborates the importance of these microRNAs for cognition and could indicate that their relation with cognition is mediated by the modulation of brain structure. For example, we determined that microRNAs were strongly associated with cortical thickness in the precentral, superior temporal (miR-10401-3p, miR-1976, miR-92b-3p, miR-4732-3p), postcentral, and middle temporal regions (miR-215-5p, miR-192-5p, miR-128-3p). The superior and middle temporal regions are implicated in a host of cognitive tasks, including audiovisual processing and integration [41], declarative memory, and language [42]. The localization in the left postcentral and precentral gyri (primary motor and somatosensory cortex) might implicate these microRNAs in normal, non-neurodegenerative brain aging, as age-related atrophy is increased in these areas [27]. Moreover, recent evidence supports the involvement of the sensorimotor region in higher cognitive functions, such as attention and learning for the primary motor cortex [43], or associative episodic memory for the somatosensory cortex [44]. Lastly, miR-340-5p and miR-125b-5p were associated with the volume of the hippocampus, perhaps the most established region in relation to memory and AD [45].

Our functional genomics analysis determined that most of the 15 microRNAs jointly regulate processes intrinsically related to cognition, such as memory, axon guidance, dendrite development, neurogenesis and synapse assembly. In addition, microRNAs from five modules jointly regulated TGF-β signaling, which plays an important role in neuronal development and neuroprotection in neurodegeneration [46,47]. Of note, this included miR-192-5p, which has been found to directly affect cognitive function through the modulation of TGF-β signaling in a mouse model of depression [48]. Despite this overlap in enrichment results, the microRNAs in each module shared few common target genes, suggesting that they are involved in different, complementary stages of the abovementioned processes. These findings point towards possible interventions for the prevention or treatment of cognitive dysfunction. For example, miR-192-5p and miR-215-5p were associated with temporal lobe thickness and episodic verbal memory. The target genes of these microRNAs were related to WNT signaling, especially known for its involvement in synaptic plasticity and AD pathology [49]. This interaction with WNT signaling, which for miR-192-5p has also been observed experimentally [50], could be further investigated as a means of preventing cortical atrophy and cognitive dysfunction in aging or AD. Besides microRNA-regulated pathways, the functional genomics analysis particularly highlighted the importance of the closely related miR-92a-3p and miR-92b-3p. A significant proportion of the target genes of these microRNAs have been linked to cognition in a previous GWAS [36]. Moreover, miR-92b-3p was associated with cortical thickness and was highly expressed in the cortex and hippocampus. Importantly, both microRNAs have been previously shown to exert neuroprotective effects [51–54].

Due to the nature of our data and analysis, we cannot conclude causality of the observed associations. However, some of the identified microRNAs have been causally linked to cognition or brain health in past experimental studies. Among them, miR-134-5p has been most extensively studied as a regulator of synaptic plasticity, dendritogenesis, long-term potentiation, and neuronal injury [55–59]. Aberrant regulation of these processes by miR-134-5p can lead to cognitive deficits [57,60,61]. Similarly, miR-409-3p has been found to induce cognitive deficits through impairment of neuronal viability [62], while miR-192-5p mediates the effects on cognitive function of exercise [63] and, as mentioned above, depression through TFG-β [48]. Moreover, miR-340-5p has been found to exert neuroprotective effects [64], while miR-493-3p, miR-1976 and miR-125b-5p worsen the neurotoxic effects of stroke [65], Parkinson’s disease [66], and AD [67].

Some of our findings were unexpected and suggest novel research questions regarding the relation of microRNAs with brain function. We identified seven microRNAs that were associated with cognition in opposite directions in our data compared to two previous studies [12,13]. We showed that this could be due to the differences in the examined populations. Specifically, for five out of seven microRNAs, the inconsistent association with cognition largely disappeared in subsets of our population that matched the age and sex composition of the previous studies. However, these inconsistent results illustrate that further validation is needed to determine the precise relation of these microRNAs with cognition. Moreover, three microRNAs (miR-128-3p, miR-92a-3p, miR-125b-5p) showed a stronger association with cognition in older participants. This could be attributed to their interaction with mechanisms of aging or neurodegeneration. For example, all three microRNAs have been implicated in AD pathogenesis [39,68,69]. Similarly, four microRNAs (miR-215-5p, miR-192-5p, miR-4732-3p, miR-340-5p) were associated with cognitive domain scores in a sex-dependent manner. This could be explained by an interaction of these microRNAs with sex hormone signaling, which is known to impact cognition [70]. In support of this, our functional enrichment results showed that miR-215-5p and miR-192-5p might regulate cellular responses to estradiol. For miR-192-5p, an interaction with estrogen receptors has also been observed experimentally [71]. However, further research is needed to define the mechanisms behind these age- and sex-dependent effects. Lastly, we observed an unexpected association of increased miR-340-5p expression with worse executive function and lower mean cortical thickness, but higher hippocampal volume. The consensus of past experimental studies supports the association of miR-340-5p with hippocampal volume, as it has been shown to have neuroprotective effects [64,72,73]. These conflicting findings could imply that this microRNA might have pleiotropic, location-dependent functions. Thus, its action might be neuroprotective in the hippocampus, but neurotoxic in the cortex, resulting in a net negative association with cognition. However, further investigation is required to test this assumption.

Our study has both strengths and limitations. As mentioned, our large sample size, wide age range, and inclusion of previous similar studies increase the reliability of our findings. Moreover, we were able to comprehensively characterize microRNAs of interest and draw conclusions as to their function, by combining an extensive neuropsychological test battery, individual-level gene expression data, detailed brain imaging measures and publicly available resources. However, our study is not without drawbacks. Our use of WGCNA is based on the assumption that similarly expressed microRNAs have shared biological functions, a principle that has been previously contested [74]. Furthermore, as is common in population-based studies, our study participants were all residents of the same geographical region, limiting the generalizability of our findings. Lastly, when comparing our results to previous similar studies, there was a relatively small overlap and even some conflicting findings. This illustrates the necessity for the validation of study results and for the standardization of microRNA sampling and measurement methods.

In conclusion, here we identified a signature of five previously identified and 10 novel microRNAs associated with cognitive function across different domains. Some of these microRNAs were concurrently related to cortical thickness and hippocampal volume, suggesting that they regulate cognitive processes through modulation of brain structure. Our functional genomics analysis demonstrated the importance of the identified microRNAs for a number of neuronal functions and emphasized miR-92a-3p and miR-92b-3p, which were abundant in cortical tissue and regulated cognition-related genes. In the future, the identified microRNAs could be leveraged to develop signatures for the early detection of dementia in clinical or research settings. Moreover, future interventional studies could utilize them for the prevention of cognitive dysfunction due to aging or neurodegeneration.

## Data Availability

Access to data can be provided to scientists in accordance with the Rhineland Study's Data Use and Access Policy. Requests for additional information or to access to the Rhineland Study's datasets can be send to RS-DUAC(at)dzne.de.

## Abbreviations

AD: Alzheimer’s Disease
ROIs: Regions Of Interest
eTIV: estimated Total Intracranial Volume
DKT: Desikan–Killiany–Tourville
SD: Standard Deviation
WGCNA: Weighted Gene Co-Expression Analysis
fdr: false discovery rate
GWAS: Genome-Wide Association Study
MCI: Mild Cognitive Impairment
CI: Confidence Interval
HCV: Hippocampal Volume
TGF-β: transforming growth factor beta

## Acknowledgements

We wish to thank all participants of the Rhineland Study and the study personnel that was involved in data collection. Additionally, we would like to thank Dr. Dan Liu for her feedback on a previous version of this manuscript.

