## Supporting information for "Circulating microRNAs are related to cognitive domains in the general population"

**Affiliations**
[a] Population Health Sciences, German Centre for Neurodegenerative Diseases (DZNE), Venusberg-Campus 1/99, 53127 Bonn, Germany
[b] AI in Medical Imaging, German Centre for Neurodegenerative Diseases (DZNE), Venusberg-Campus 1/99, 53127 Bonn, Germany
[c] Department for Epigenetics and Systems Medicine in Neurodegenerative Diseases, German Center for Neurodegenerative Diseases, Von-Siebold-Str. 3a, 37075 Göttingen, Germany
[d] Bioinformatics Unit, German Centre for Neurodegenerative Diseases, Von-Siebold-Str. 3a, 37075 Göttingen, Germany
[e] Department of Neurology, Faculty of Medicine, University of Bonn, Venusberg-Campus 1/80, 53127 Bonn, Germany
[f] Department for Psychiatry and Psychotherapy, University Medical Center Göttingen, Von-Siebold-Str. 5, 37075 Göttingen, Germany
[g] Cluster of Excellence MBExC, University of Göttingen & University Medical Center Goettingen, Robert-Koch-Str. 40, 37075 Göttingen, Germany
[h] Institute for Medical Biometry, Informatics and Epidemiology (IMBIE), Faculty of Medicine, University of Bonn, Venusberg-Campus 1/11, 53127 Bonn, Germany

### Supplementary materials

**Supplementary Methods S1:** MicroRNA and gene expression sequencing and analysis
**Supplementary Methods S2:** Weighted gene co-expression network analysis
**Supplementary Methods S3:** MicroRNA target genes and functional enrichment analysis
**Supplementary Table S1:** Previous studies that identified circulating microRNAs related to cognition, AD or MCI
**Supplementary Figure S1:** Construction of weighted gene co-expression network
**Supplementary Figure S2:** Association of microRNA expression with cognitive scores in Model 2.
**Supplementary Figure S3:** Distribution of normalized counts for cognition-related microRNAs
**Supplementary Figure S4:** Neuropsychiatric and neurodegenerative disease sensitivity analysis.
**Supplementary Figure S5:** Stratified analysis for microRNAs inconsistently associated with cognition in our study and in two previous studies
**Supplementary Figure S6:** Predicted and downregulated microRNA target genes
**References**

### Supplementary Methods 1

*MicroRNA sequencing and analysis*

Sequencing libraries for microRNA were prepared using NEBNext® small RNA library preparation kit according to manufacturer´s instructions. Quality control of the sequencing was evaluated through FastQC v0.11.9 [1]. Trimmomatic v.0.39 software [2] was used to remove sequencing adapters, low-quality score reads, and reads shorter than 18 base pairs. The alignment and the quantification of microRNAs was performed using the mirdeep2 version 2.0.1.2 software package [3]. Briefly, following the trimming of low-quality score reads, the sequencing reads were aligned to the Human Genome GRCh38.p13 provided by Ensembl, and the expression of known microRNAs was determined by mapping the sequencing reads to predefined human precursor and mature microRNA sequences retrieved from miRBase v22.1 [4].

*Gene expression sequencing and analysis*

RNA integrity and quantity were assessed through the tapestation RNA assay on a Tapestation4200 instrument (Agilent). 750 ng of total RNA was used to generate NGS libraries for total RNA sequencing using the TruSeq stranded total RNA kit (Illumina) following manufacturer's instructions with Ribo-Zero Globin reduction. We checked library size distribution using D1000 on a Tapestation4200 instrument (Agilent) and quantified the libraries via Qubit HS dsDNA assays (Invitrogen). We clustered the libraries at 250 pM final clustering concentration on the NovaSeq6000 using S2 v1 chemistry (Illumina) in XP mode and sequenced paired-end 2*50 cycles before demultiplexing using bcl2fastq2 v2.20. Quality control of the sequencing was evaluated through FastQC v0.11.9. Following the trimming of low-quality score reads using Trimmomatic v.0.39 software, sequencing reads were aligned to the human reference genome GRCh38.p13 from Ensembl using STAR v2.7.1 [5]. The count matrix was generated with STAR –quantMode GeneCounts using the human gene annotation version GRCh38.101.

### Supplementary methods 2

*Construction of weighted gene co-expression network*

To construct the WGCNA dendrogram, we first calculated pair-wise biweight midcorrelations of microRNA expression. We then chose a soft thresholding power of 7, which satisfied the scale-free topology criterion of $R^2>0.9$ (**Supplementary Figure S1**), and used a signed network to construct an adjacency matrix. Next, we converted the adjacency matrix to a topological matrix, using the topological overlap measure (TOM), and generated a hierarchical clustering tree with 1-TOM as the distance and using the UPGMA algorithm (method=”average”). Using the Hybrid Dynamic Tree Cut method and setting a minimum module size of 5, we thus identified 16 microRNA modules. We set the *deepsplit* parameter to 4, to increase the overall number of clusters and avoid over-clustering.

*Identification of module eigengenes and hub microRNAs*

WGCNA permits users to reduce the expression of genes in each module to a singule value. This value, called the module (eigen)expression, is defined as the first principal component derived from Principal Component Analysis (PCA), and is representative of overall microRNA expression within the module. Using this method, we calculated the eigenexpression of each of the microRNA modules identified in our data. Another useful tool offered by WGCNA is the identification of *hub genes* within each module, which are seen as module representatives, or key drivers of module expression and its relation to traits. We defined *hub microRNAs* as microRNAs with *high module membership* and *high significance*, that is, microRNA expression had to be *a)* highly correlated with module eigenexpression (Pearson's R ≥ 0.8 between microRNA expression and module eigenexpression) and *b*) significantly associated with the same trait as module expression.

**Supplementary Methods 3**

*Identification of microRNA target genes*

To identify genes that could be regulated by microRNAs, we examined the association between the expression of microRNAs and their potential target genes. We utilized the *multimir* R package (v.1.12.0) [6] to obtain a list of putative target genes for each microRNA from three online databases: experimentally validated microRNA-target gene interactions were obtained from the MirTarBase [7] database, while predicted microRNA-target gene interactions were obtained from the TargetScan [8] and miRDB [9,10] databases. To ensure the inclusion of all putative microRNA-target genes, the databases were interrogated for all predicted target genes, regardless of database prediction scores, and we employed the union of the target genes obtained from the 3 databases. Subsequently, we constructed separate regression models for each microRNA (independent variable) and their putative target genes (dependent variable), adjusting for age, sex, and blood cell counts. Lastly, we filtered for negatively associated target genes (linear regression beta coefficient<0 and *P* value ≤ .05), based on the assumption that microRNAs would downregulate the expression of their target genes.

*Functional enrichment analysis*

For the identified target genes we performed a pathway enrichment analysis using the *clusterProfiler* (v. 3.18.1) R Bioconductor package [11]. As we assume that microRNAs in WGCNA modules jointly regulate biological functions, we pooled the target genes of all the microRNAs belonging to the same module. We then conducted an overrepresentation analysis per module, using enrichment terms obtained from the Gene Ontology: Biological Processes (Gene Ontology) [12] database. The universe (background genes) of the overrepresentation analysis was set to the genes measured in our study (n=11,325). As enrichment terms from Gene Ontology tend to be redundant, similar terms were collapsed using the *rrvgo* (v. 1.2.0) R Bioconductor package [13]. Specifically, a semantic similarity matrix of Gene Ontology terms was constructed and similar terms were grouped, selecting the largest term (as defined by the number of genes in each enriched term) to represent each group. A small similarity threshold of 0.6 was selected to group only highly redundant terms, and the rest of the parameters were left at default settings. The *P* values of the terms within each group were pooled using the Fisher method and multiple testing adjustment according to the Benjamini-Hochberg false discovery rate method was performed for the pooled *P* values.

**Supplementary Table S1: Previous studies that identified circulating microRNAs related to cognition, AD or MCI**

| Study title | Study type | Sample size | MicroRNAs identified* | Reference |
|---|---|---|---|---|
| Genome-wide profiling of circulatory microRNAs associated with cognition and dementia. | Population-based, cross-sectional | n=1615 | miR-150-5p, miR-19b-3p, miR-20b-5p, miR-122-5p, miR-92a-3p, miR-4732-3p, miR-146a-5p, miR-30a-5p, miR-23b-3p, let-7c-5p, miR-4449, miR-324-3p , miR-6780b-5p, miR-2861, miR-4800-5p , miR-4688, miR-6800-3p , miR-185-5p , miR-3155b, miR-372-3p, miR-6126, miR-8071, miR-4697-5p, miR-7106-5p, miR-187-3p, miR-1322, miR-1275, miR-3141, miR-8078, miR-197-5p, miR-6870-5p, miR-6738-5p, miR-4539, miR-4522, miR-34a-5p, miR-4534 (Identified in multi-marker analysis) | Yaqub et al, 2022 [15] |
| Extracellular microRNA and cognitive function in a prospective cohort of older men: The Veterans Affairs Normative Aging Study. | Population-based (male participants), cross-sectional and longitudinal | n=530 | Cross-sectional analysis: miR-148a-5p, miR-335-3p<br>Longitudinal analysis: miR-28-3p, miR-152-3p, miR-30e-3p, miR-148b-3p, miR-326, miR-22-3p, miR-221-3p, miR-4732-3p, miR-652-3p, let-7b-5p, miR-342-5p, miR-574-5p, miR-21-5p, miR-320c, miR-340-5p, miR-194-5p, miR-6842-3p, miR-363-3p, miR-584-5p, miR-99a-5p, miR-191-5p, miR-329-3p, miR-215-5p, miR-181a-5p, let-7f-5p, miR-320b, miR-199a-5p, miR-125b-5p, miR-511-5p, miR-6852-5p, miR-431-5p, miR-493-5p, miR-337-3p | Comfort et al, 2022 [14] |
| A microRNA signature that correlates with cognition and is a target against cognitive decline. | Population-based, mouse models, case-control | Population-based: n=132; Case-control: n=71 MCI patients, n=65 HC | miR-181a-5p, miR-146a-5p, miR-148a-3p | Islam et al, 2021 [16] |
| The interplay between micro RNAs and genetic liability to Alzheimer's Disease on memory trajectories in the general population. | Population-based, longitudinal | n = 548 | miR-19a-3p, miR-485-5p | Van der Auwera et al, 2023 [17] |
| Association of Blood MicroRNA Expression and Polymorphisms with Cognitive and Biomarker Changes in Older Adults. | Population-based, cross-sectional | n=830 | let-7a-5p, let-7c-5p, let-7d-5p, miR-144-5p, miR-93-5p, miR-98-5p, miR-363-3p (Identified in linear regression analysis) | Sadlon et al [18] |
| Differential expression of MicroRNAs in Alzheimer's disease: a systematic review and meta-analysis | Meta-analysis | AD: n=2,787 patients, n=2,726 HC; MCI: n=302 patients, n=408 HC | AD: miR-103a-3p, miR-106a-5p, miR-107, miR-1306-5p, miR-146a-5p, miR-15a-5p, miR-15b-3p, miR-20a-5p, miR-27b-3p, miR-29b-3p, miR-30e-5p, miR-335-5p, miR-361-5p, miR-18b-5p, has-miR-221-3p, miR-31-5p, miR-424-5p, miR-582-5p, miR-9-5p<br>MCI: miR-128-3p, miR-132-3p, miR-323a-3p, miR-134-3p, miR-874-3p (microRNAs differentially expressed in blood with $I^2<0.4$) | Yoon et al, 2022 [19] |

* Underlined microRNAs were measured in the Rhineland Study.
Abbreviations: MCI, Mild Cognitive Impairment; HC, Healthy Control; AD, Alzheimer's Disease

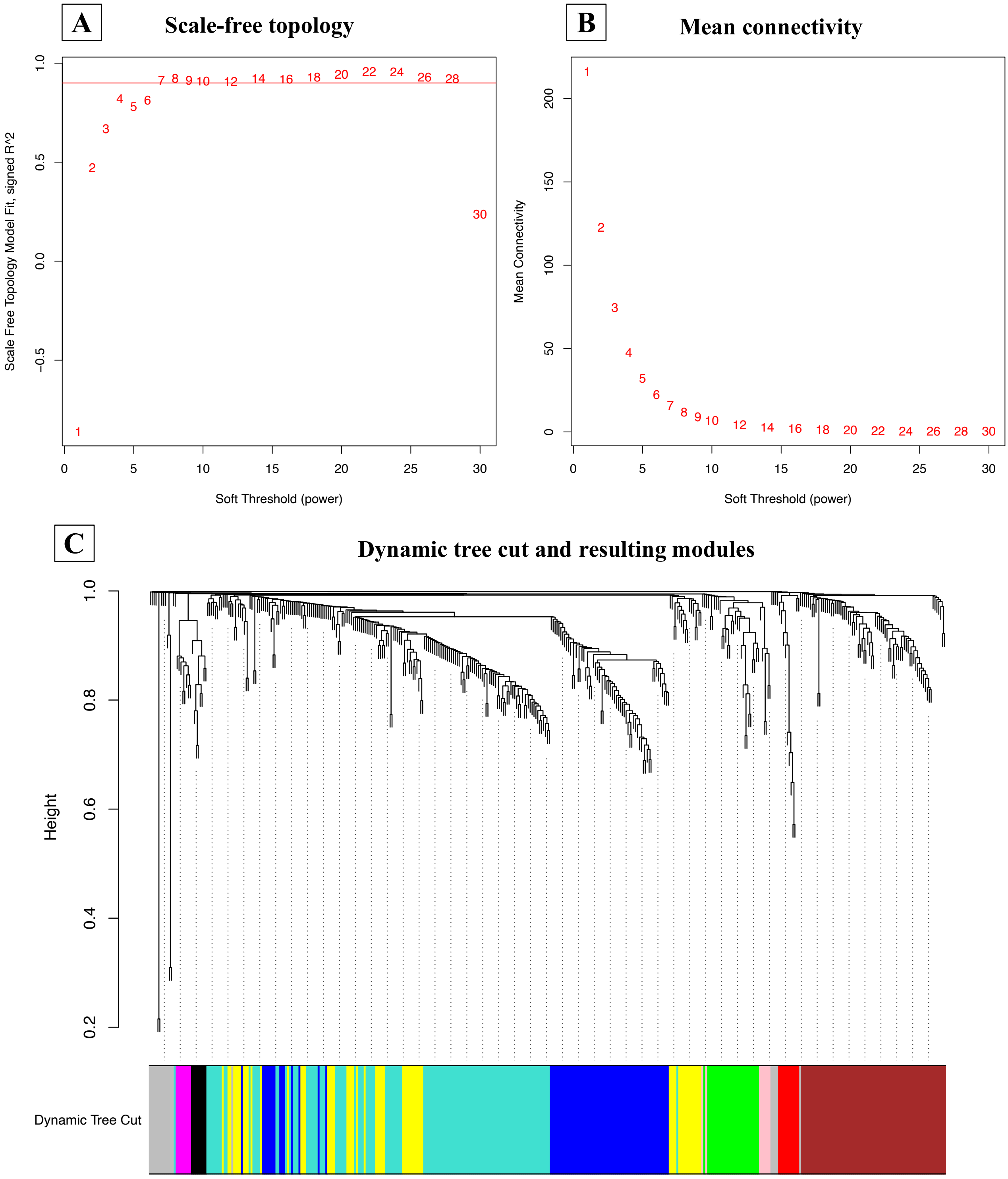


**Supplementary Figure S1**: Construction of weighted gene co-expression network
**(A)** Scale-free topology and (**B)** mean connectivity as a function of soft thresholding power.
**(C)** Implementation of the dynamic tree cut algorithm and distribution of microRNAs in color-coded clusters (modules). Resulting modules are shown as colors at the lower part of the plot.

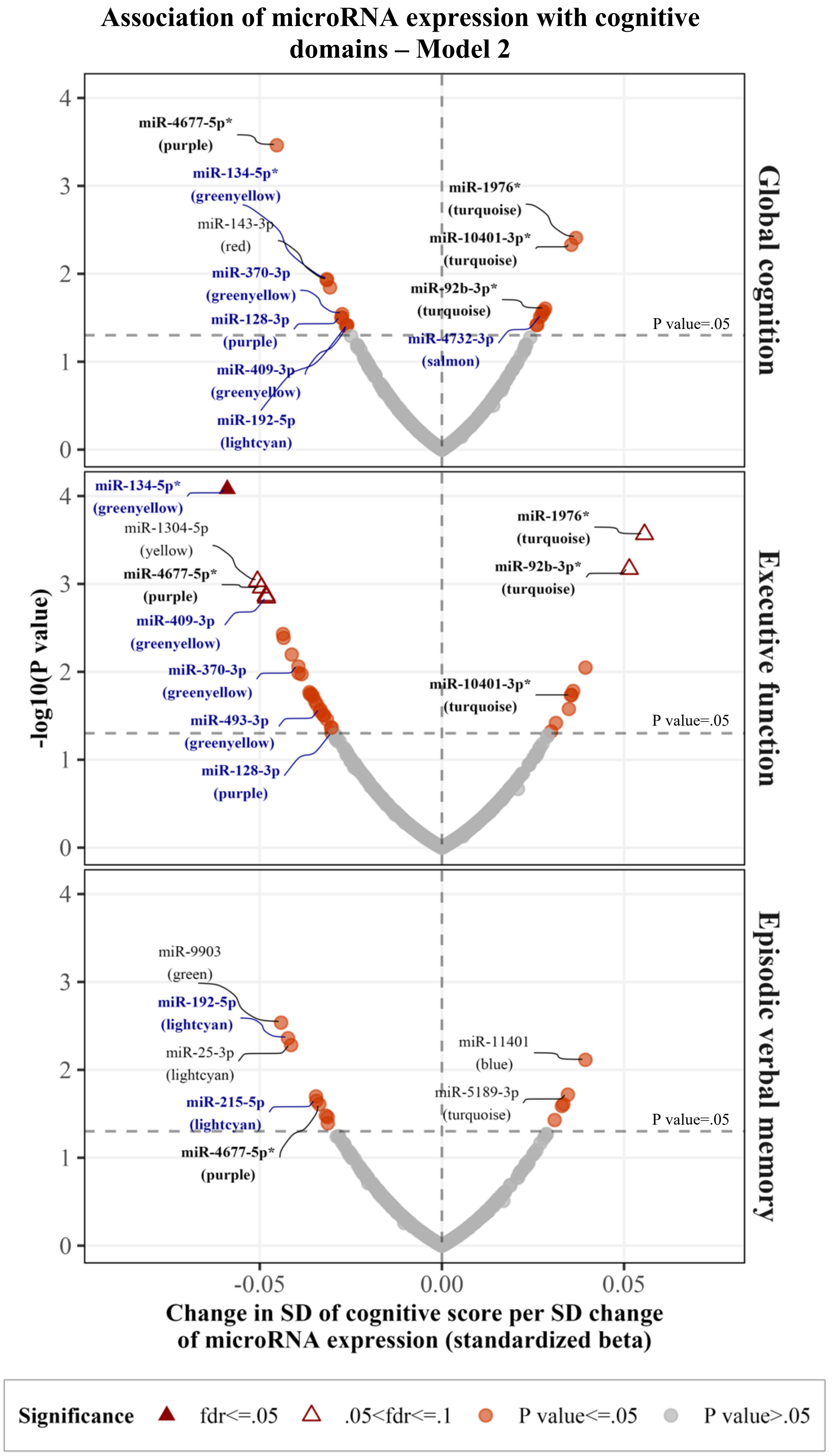


**Supplementary Figure S2:** Association of microRNA expression with cognitive scores in Model 2. Volcano plots of the association of all microRNAs profiled in our study with global cognition, executive function and episodic verbal memory scores. The *P* value shown in the y-axis has not been adjusted for multiple testing. Top microRNAs for each score are annotated and co-expression modules are shown in parentheses. MicroRNAs that reached statistical significance for at least one phenotype, after multiple testing correction (fdr ≤ 0.05), are shown with triangles and have been marked with an asterisk (*). Hub microRNAs are annotated with blue color. Model 2 was used, which included as covariates age, sex, first language and educational level.
Abbreviations: SD, Standard Deviation, fdr, false discovery rate

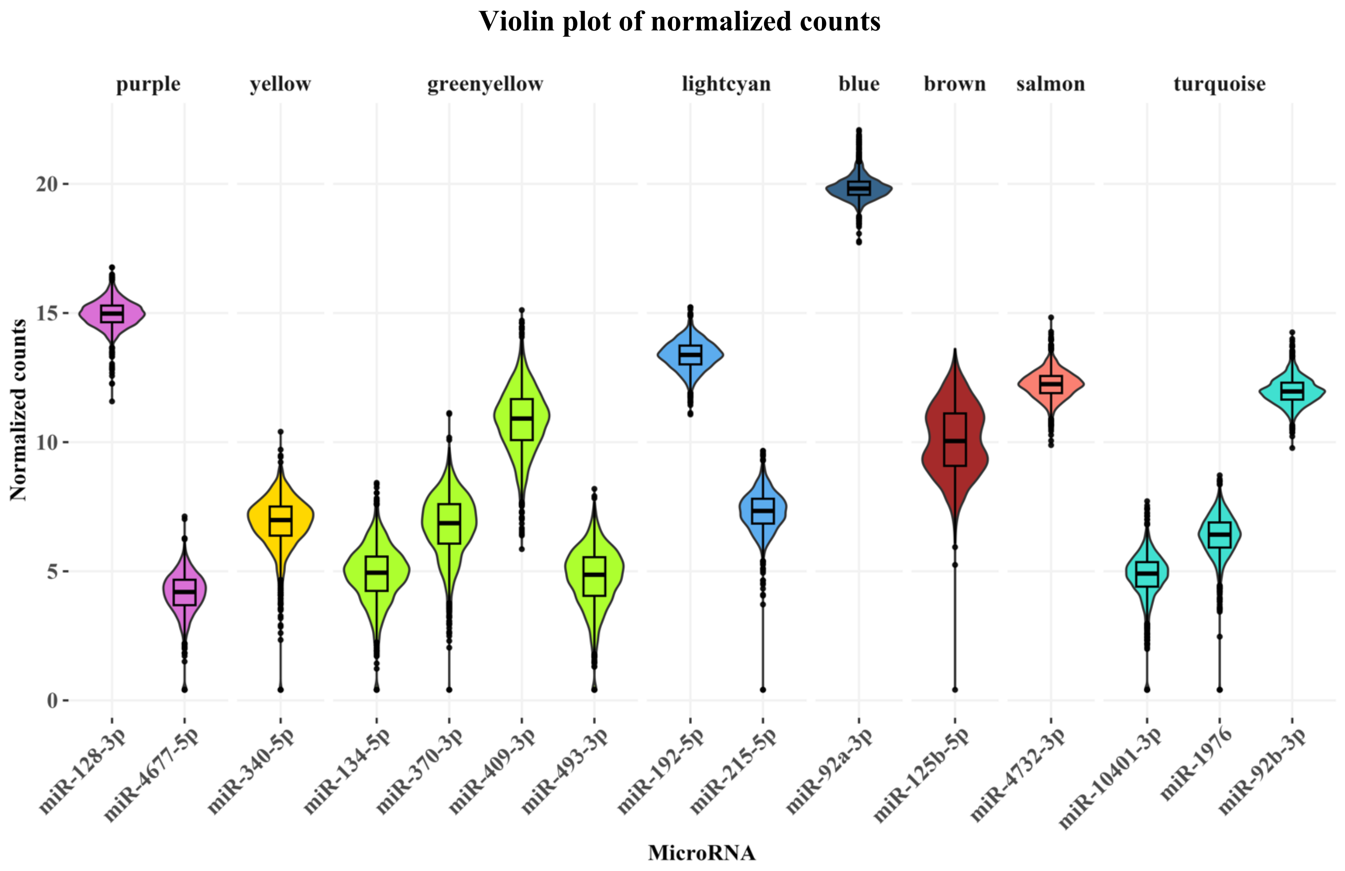


**Supplementary Figure S3:** Distribution of normalized counts for cognition-related microRNAs The boxplots and violin plots show the distribution of DESeq2-normalized counts for the 15 identified cognition-related microRNAs, before adjusting for sequencing batch. MicroRNAs have been grouped according to the Weighted Gene Co-expression module they belong to.

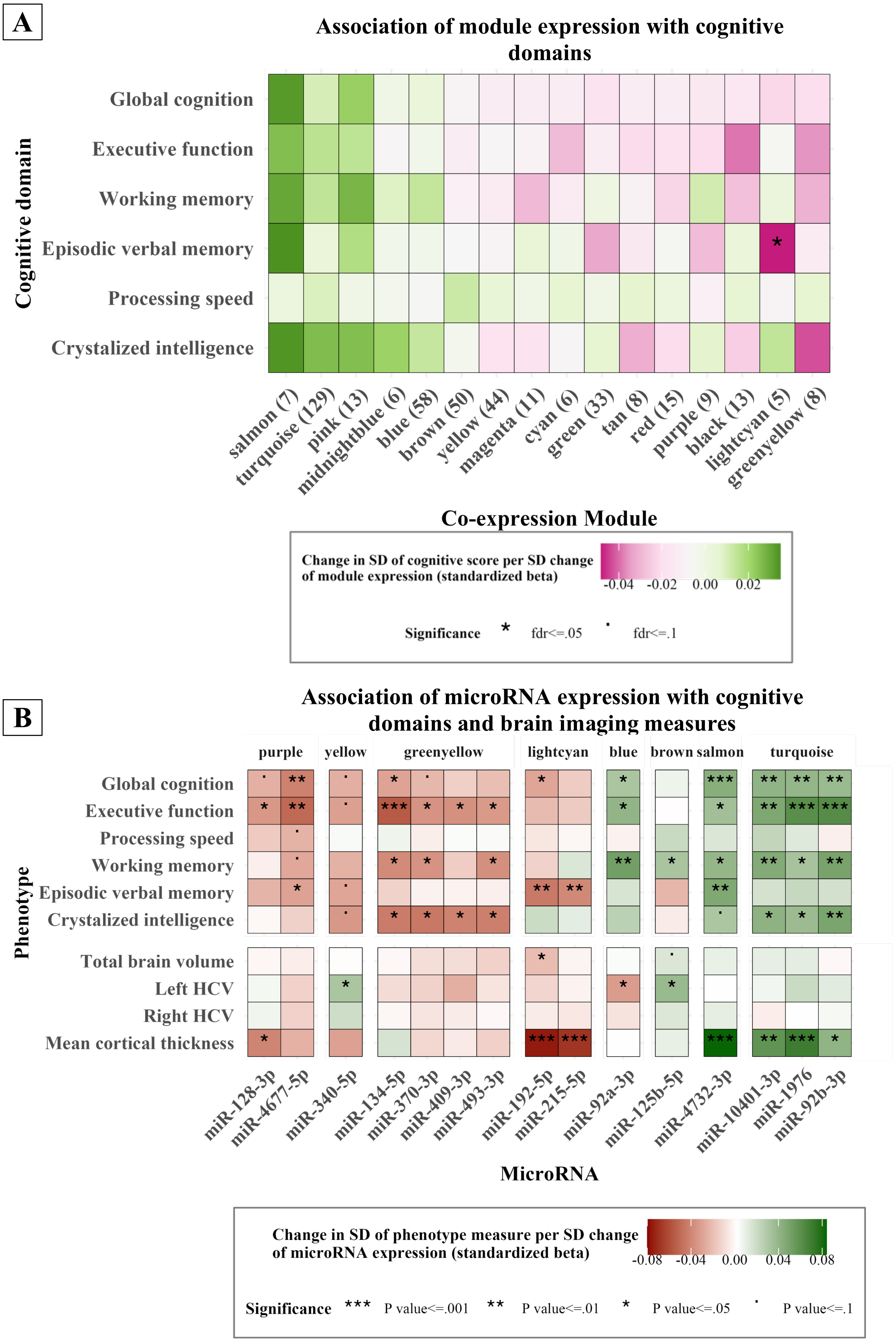


**Supplementary Figure S4:** Neuropsychiatric and neurodegenerative disease sensitivity analysis. Linear regression coefficients and significance levels for the association of (**A**) co-expression modules and (**B**) individual microRNAs with cognitive scores, after exclusion of participants who reported a physician diagnosis of dementia, Parkinson’s disease, multiple sclerosis, stroke, and schizophrenia.

**A**

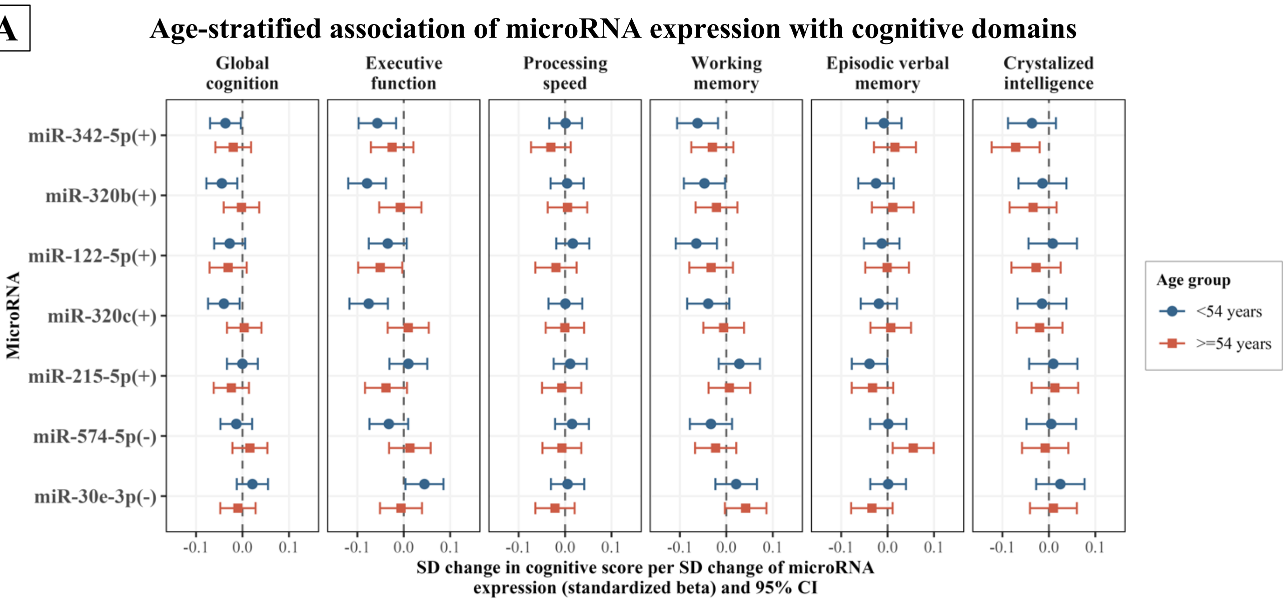


**B**

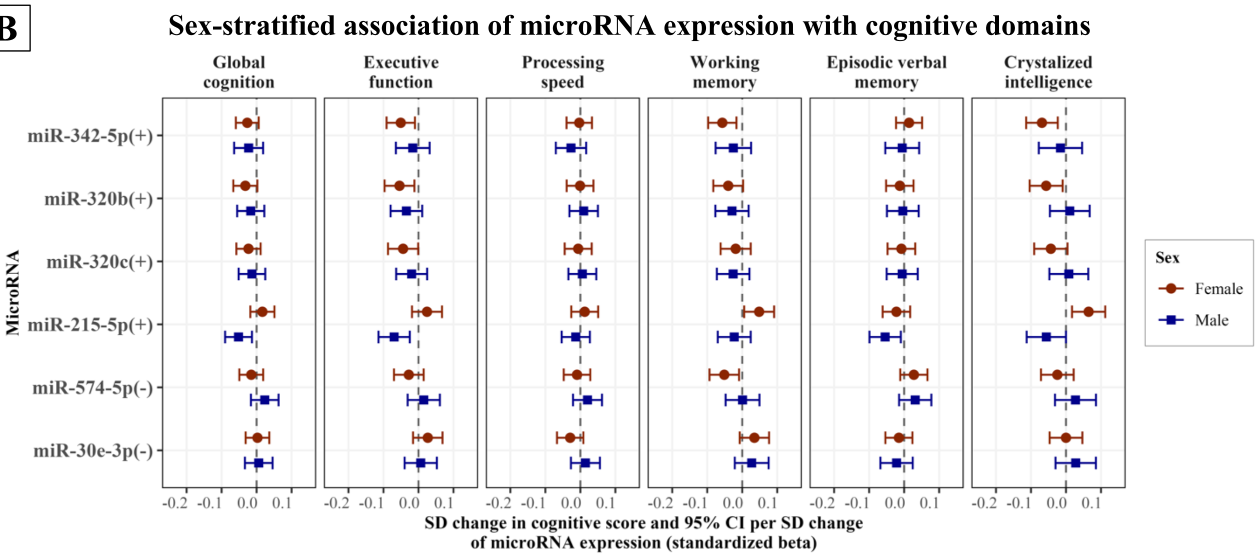


**Supplementary Figure S5:** Stratified analysis for microRNAs inconsistently associated with cognition in our study and in previous studies.
Seven microRNAs were associated with cognition in a different direction in our study and in two previous studies [14,15]. The forest plot shows the standardized regression betas and 95% confidence intervals the association of these microRNAs with cognitive scores after stratifying by **(A)** age and (**B**) sex. MicroRNAs are marked with “+” when their higher expression was associated with better cognitive function in the previous studies, and with “-” in the opposite case. Linear regression analyses were based on Model 1, which was adjusted for age.
Abbreviations: SD, Standard Deviation; CI, Confidence Interval

**A**

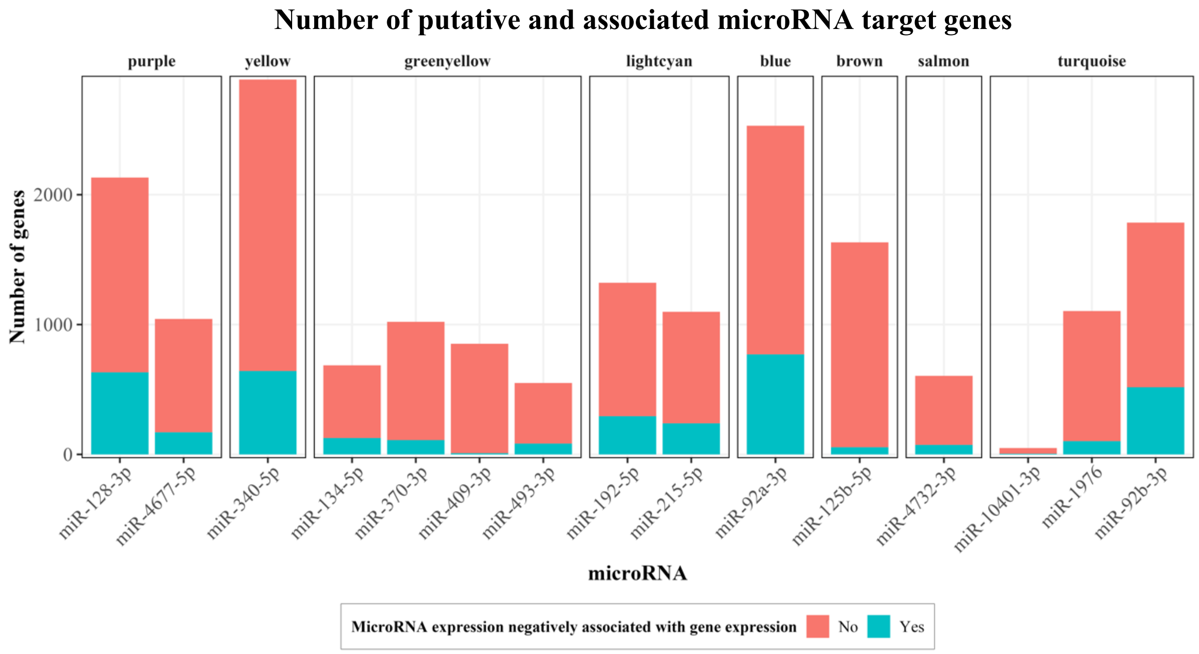


**B**

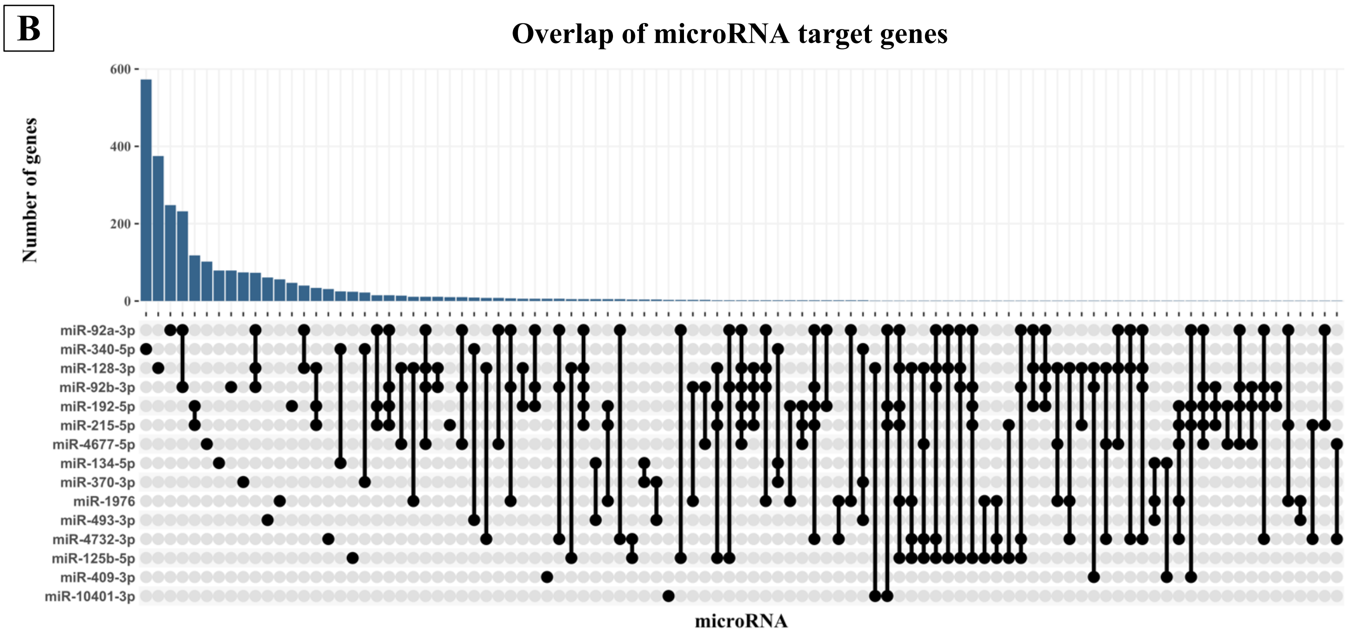


**Supplementary Figure S6:** Predicted and downregulated microRNA target genes. (**A**) The bar plot shows the number of genes that were predicted to be targeted by microRNAs, and the proportion of these predicted targets the expression of which was negatively associated with the expression of the targeting microRNA. MicroRNAs have been grouped by Weighted Gene Co-expression module. (**B**) The upset plot shows the overlap of negatively associated target genes per microRNA. The microRNAs are represented by dots. The lines linking the dots denote that the corresponding microRNAs have common negatively associated target genes. The number of these genes is shown in the bar plot above.
